# Risk prediction for lung cancer screening: a systematic review and meta-regression

**DOI:** 10.1101/2025.09.10.25335529

**Authors:** Ramin Rezaeianzadeh, Crystal leung, Soo Jeong Kim, Kayly Choy, Kate Johnson, Miranda Kirby, Stephen Lam, Benjamin Smith, Mohsen Sadatsafavi

## Abstract

**Background:** Lung cancer (LC) is the leading cause of cancer mortality, often diagnosed at advanced stages. Screening reduces mortality in high-risk individuals, but its efficiency can improve with pre- and post-screening risk stratification. With recent LC screening guideline updates in Europe and the US, numerous novel risk prediction models have emerged since the last systematic review of such models. We reviewed risk-based models for selecting candidates for CT screening, and post-CT stratification.

**Methods:** We systematically reviewed Embase and MEDLINE (2020–2024), identifying studies proposing new LC risk models for screening selection or nodule classification. Data extraction included study design, population, model type, risk horizon, and internal/external validation metrics. In addition, we performed an exploratory meta-regression of AUCs to assess whether sample size, model class, validation type, and biomarker use were associated with discrimination.

**Results:** Of 1987 records, 68 were included: 41 models were for screening selection (20 without biomarkers, 21 with), and 27 for nodule classification. Regression-based models predominated, though machine learning and deep learning approaches were increasingly common. Discrimination ranged from moderate (AUC≈0.70) to excellent (>0.90), with biomarker and imaging-enhanced models often outperforming traditional ones. Model calibration was inconsistently reported, and fewer than half underwent external validation. Meta-regression suggested that, among pre-screening models, larger sample sizes were modestly associated with higher AUC.

**Conclusion:** 75 models had been identified prior to 2020, we found 68 models since. This reflects growing interest in personalized LC screening. While many demonstrate strong discrimination, inconsistent calibration and limited external validation hinder clinical adoption. Future efforts should prioritize improving existing models rather than developing new ones, transparent evaluation, cost-effectiveness analysis, and real-world implementation.

## Introduction

Lung cancer remains one of the most common, and one of the deadliest, malignancies worldwide, accounting for an estimated 11.6 percent of all new cancer diagnoses and 18.4 percent of cancer related deaths in 2018.^1^ Diagnosis at an advanced stage drives dismal outcomes, with a five year survival of only about 3 percent, whereas detection at an early stage improves five year survival to approximately 62 percent.^2^ Most lung cancer cases are diagnosed at later stages. Almost half (44.8%) diagnosed lung cancer cases from 2017 to 2021 were already spread to distant parts of the body.^34^ Screening for lung cancer offers the possibility to shift the diagnosis stage towards earlier, more curable stages of its progression.^5^

Large randomized trials have established low-dose computed tomography (LDCT) as the preferred screening tool for individuals at high risk. ^6–13^ In particular, the U.S. National Lung Screening Trial (NLST) demonstrated a 20 percent relative reduction in lung cancer mortality with LDCT versus chest radiography.^14^ Accordingly, major guideline bodies now recommend annual LDCT screening for individuals exceeding certain smoking levels. In 2021, the U.S. Preventive Services Task Force (USPSTF) expanded its recommendations for screening to cover adults aged 50–80 years with a ≥ 20 pack-year smoking history who currently smoke or quit within the past 15 years.^15^

Parallel efforts in Europe and elsewhere have followed: after trials such as NELSON^11^ and MILD^8^, countries including Croatia^16^ (nationwide since 2020), the Czech Republic (2022)^17^, Poland^18^ (2021–22), and the United Kingdom^18^ (2023) have adopted LDCT screening programs. In 2022, the European Commission urged all 27 EU member states to implement a “stepwise” approach to lung cancer screening.^19^ Canada’s Task Force on Preventive Health Care similarly recommends screening adults aged 55–74 years with a ≥ 30 pack-year history for up to three consecutive years.^20^

Despite these advances, current eligibility criteria still include many who will never develop lung cancer and exclude a substantial proportion of those who ultimately die from it.^21,22^ To improve the efficiency of screening, risk-based strategies have been proposed that incorporate additional clinical, demographic, and radiologic factors. Such risk-based strategies are generally applied for two purposes: firstly, to identify individuals at highest baseline risk who stand to benefit most from LDCT screening, and secondly, to stratify malignancy risk in persons with pulmonary nodules detected on LDCT.

There are no universally accepted risk scoring algorithms, and developing new algorithms is an active area of research. A 2020 systematic review identified 75 novel risk prediction models published until 2020.^23^ Perhaps due to recent policy updates (e.g., the 2021 USPSTF guidelines), evolving regulatory frameworks in Europe and Asia, and the emergence of machine-learning– driven approaches, several new models have been proposed since the publication of the aforementioned systematic review.

In light of the above, we have conducted an updated appraisal. The purpose of this work was to identify and describe novel risk prediction models developed across diverse jurisdictions since 2020, with a focus on appraising statistical methodology, incorporation of novel biomarkers, and various statistical strategies for predicting the risk of lung cancer among high-risk individuals.

## Methods

We conducted a systematic literature search to identify relevant studies published between January 2020 and November 2024. This review updates a previous systematic review on risk-based lung cancer screening conducted by Toumazis et al. published in 2020.^23^ The search strategy, developed in consultation with an in-house librarian, combined both controlled vocabulary and free-text terms, including “lung cancer screening”, “risk prediction”, “risk-based screening”, and “risk assessment”, (Supplementary Table 1). We developed and documented our search strategy prior to execution, to ensure comprehensive retrieval of literature, across the databases used. Studies were eligible for inclusion if they met the following criteria: (1) the study proposed a novel lung cancer prediction model, (2) the full-text article was obtainable, and (3) the manuscript was written in English. Articles were excluded if any of the following applied: (1) the record was a review article, editorial, letter, or commentary, (2) the study was not published in a peer-reviewed journal, (3) the study was not conducted in a screening setting (such as studies involving only full-dose CT scans or invasive tissue collection for model training), or (4) the study examined lung cancer risk in populations with an existing underlying condition.

The search was conducted in the Ovid Embase and Medline databases. We screened results in two stages. In the first stage, titles and abstracts were independently reviewed by two reviewers (RR and CL). In cases of disagreement, a third reviewer (MS) was consulted to resolve conflicts, ensuring a rigorous and unbiased study selection process. In the subsequent stage, potentially eligible studies underwent a full-text review using the same process for resolving discrepancies. Data extraction was carried out by one reviewer (RR) using an adapted version of a previously published extraction form^24^, and the extracted data were subsequently reviewed by two additional reviewers (CL and KC) for accuracy and consistency. Information extracted from each study included details regarding: (1) the risk factors incorporated into each lung cancer prediction model, (2) the study design, (3) the model type, (4) the targeted population, (5) the prediction type, and (6) key performance metrics. The performance metrics included measures of accuracy (sensitivity, specificity, predictive values at reported threshold values), calibration measures assessing the agreement between predicted probabilities and observed outcomes, and discrimination measures such as the area under the receiver operating characteristic (ROC) curve. Performance measures were only interpreted if they were from an external validation (assessed in a sample not used for training the model) or from an internal validation (based on the same data used to fit the model) after optimism correction (e.g., via bootstrapping or cross validation). We considered uncorrected estimates of model performance from internal validation uninterpretable.

The data were summarized in tabulated form with accompanying narrative summaries to enable direct comparisons of predictive performance across models. We categorized models by the screening step they address, that is risk-based lung cancer screening models used to determine screening eligibility, without biomarkers and with biomarker data. Or post-screening malignancy risk prediction models for screen-detected pulmonary nodules.

### Meta-regression

We conducted an exploratory meta-regression to explore heterogeneity among the included studies on estimates of discrimination. We extracted study-level data on area under the curve (AUC, or c-statistic) values with their standard errors (SE). For each AUC estimate, we recorded the following variables: sample size, prediction model category (regression [baseline]; machine-learning approaches; neural networks; all else were categorized as other), validation type (internal [baseline]; external), and, in pre–screening studies, whether biomarkers were included in the model.

If a study reported multiple AUCs for different subgroups (e.g., in different validation samples), we included all of them as long as the samples did not overlap. To account for clustering of AUCs within each study, we conducted meta-regressions with robust variance estimation using the *robumeta* package. Given heterogeneity in model specifications and the time points at which AUC was measured, we did not report pooled AUC estimates by screening stage. All analyses were performed in R (version 4.4.2).

## Results

Of the 1,987 records identified through database searching (1,344 from Embase and 643 from MEDLINE), 379 duplicates (one manually and 378 by Covidence) were removed, leaving 1,608 unique records for title and abstract screening. Of these, 1,389 were excluded, and the remaining 217 articles underwent full-text retrieval (all were successfully retrieved). Full-text assessment led to exclusion of 148 studies. 68 studies met inclusion criteria and were incorporated into the review (Figure 1). These studies reported on 41 new models on risk prediction models before screening, of which 21 included a biomarker. 27 new models were developed to predict the risk of malignancy in identified lung nodules after screening.

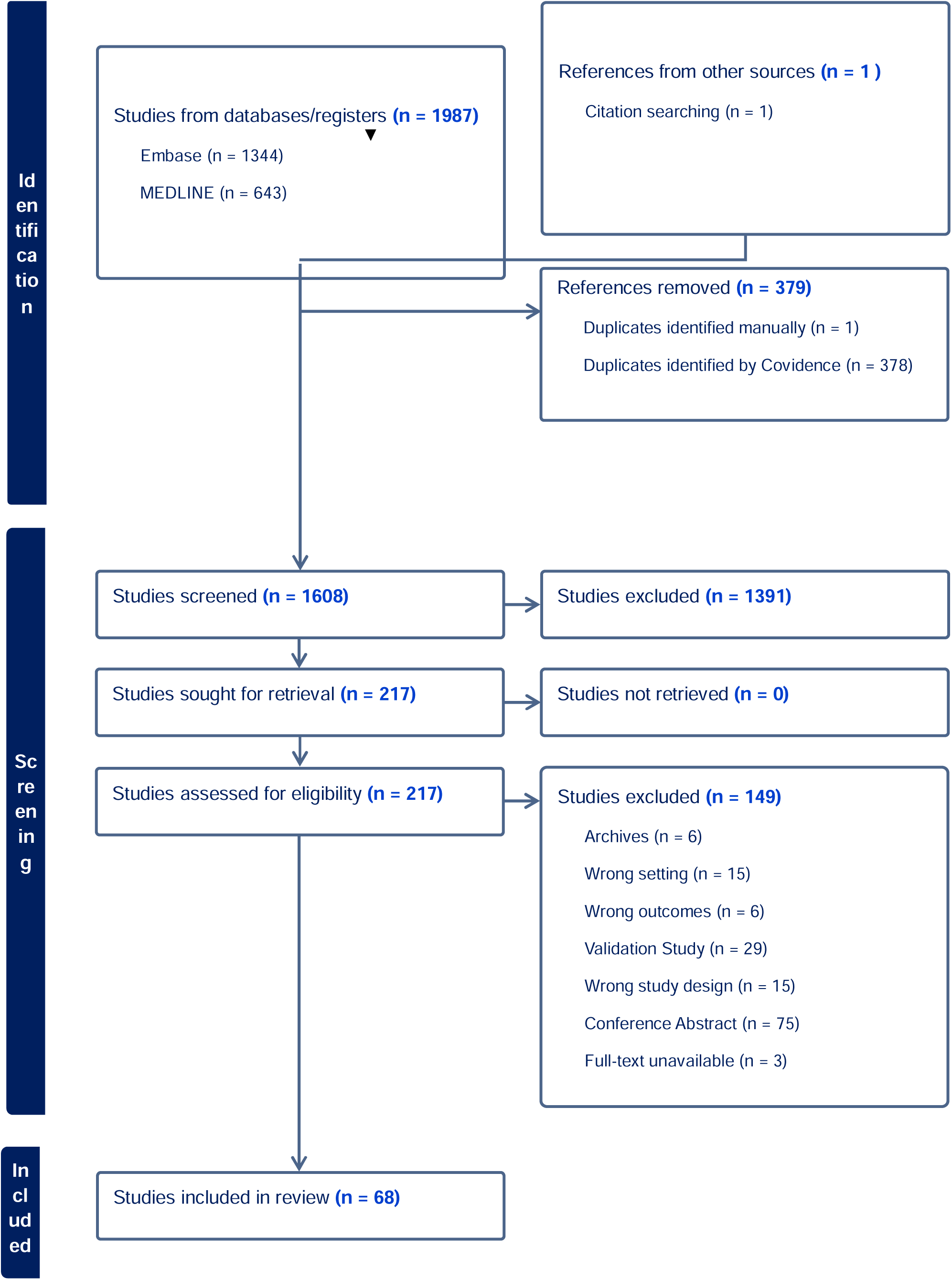

***Table 1*** provides the list of new risk prediction models for lung cancer risk for screening selection without incorporating a biomarker. The left-most column provides a shorthand label for each model to facilitate referral to the table in the text below.

**Table 1.** Risk-based lung cancer screening models not incorporating biomarkers.

|  |  |  |  |  |  |  |  |  |  |  |  |  |
| --- | --- | --- | --- | --- | --- | --- | --- | --- | --- | --- | --- | --- |
| Study Number | Risk Prediction Model | Risk Factors | Study Design | Model Type | Targeted Population | Prediction Type | Predictive Performance |  |  |  | Sensitivity (threshold) | Specificity (threshold) |
|  |  |  |  |  |  |  | Internal Validation |  | External Validation |  |  |  |
|  |  |  |  |  |  |  | Calibration | Discrimination | Calibration | Discrimination |  |  |
| LCR001 | Shanghai-LCM<sup>25</sup> | Age; sex; education; smoking status; smoking years; pack-years; years quit; BMI; history of lung diseases; family history | Prospective | Cox proportional hazards | Asian ever-smokers | Lung cancer incidence within 1–10 years | Slope: Original 1.0000; Training 1.0000; Test 0.9839; Optimism 0.0161; Index-corrected 0.9839; E/O 0.70 (95% CI) | C-statistic: Original 0.7805; Training 0.7826; Test 0.7787; Corrected 0.7765<br>Somers' Dxy: Original 0.5609; Training 0.5652; Test 0.5573; Corrected 0.0079 | – | C-statistic 0.70 (95% CI 0.67–0.72) | – |  |
| LCR002 | Levi<sup>26</sup> | Age; gender; socioeconomic status; BMI; smoking history; COPD/emphysema/chronic bronchitis; ILD/pulmonary fibrosis; family history | Retrospective | Ensemble ML (Random Forest + XGBoost) | General population | Lung cancer incidence (medical records) | – | Accuracy: Overall 71.2%; Smokers 74.8%;<br>Non-smokers 73%<br>F1-score 71.4%; PPV 74% | – | – | 69% (overall) | – |
| LCR00 | Guo<sup>27</sup> | Age; gender; | Prospective | Cox | Urban | Lung | Assesse | C-statistic: | – | – | – | – |
| 3 |  | smoking pack-years; history of tuberculosis; history of emphysema | ve | proportional hazards | Chinese aged 40–74 | cancer diagnosis at 1-, 3-, 5-year intervals | d<br>graphically | Training<br>1-yr 0.766;<br>3-yr 0.739;<br>5-yr 0.745<br>Validation<br>1-yr 0.741;<br>3-yr 0.734; 5-yr 0.735<br>Subgroups<br>(male/female;<br><60/≥60;<br>never/current/former) ranged 0.591–0.791 |  |  |  |  |
| LCR005 | OWL<sup>28</sup> | Demographic (age, gender, BMI, education), smoking (status, age at beginning smoke, average numbers of cigarettes per day, duration of smoking, durations since quitting, pack-years), clinical history (diabetes, COPD, emphysema, chronic bronchitis), and family history of lung cancer | Prospective | XGBoost | UK Biobank, PLCO, NLST cohorts | 5-year invasive lung cancer diagnosis | E/O 5-yr 1.002 (95% CI 0.899–1.133);<br>6-yr 1.037 (95% CI 0.939–1.159) | C-index 0.86 (95% CI 0.85–0.87);<br>5-fold CV: 0.84 | – | C-index Across PLCO & NLST: 0.86 (95% CI 0.84–0.87); 0.84 (95% CI 0.83–0.85) | – | – |
| LCR00 | CanPre | Age; sex; ethnicity; | Retrospec | Cox | Genera | Lung | Assesse | C-statistic: | Assesse | C-statistic: | – | – |
| 7 | dict<sup>29</sup> | Townsend score; smoking status/intensity; alcohol; BMI; COPD; asthma; pneumonia; VTE; asbestos; family history; personal cancer history; age × smoking interaction | tive | proportional hazards | l population | cancer incidence up to 10 years | d graphically | WOMEN: 0.897 (0.893-0.900), MEN: 0.904 (0.901-0.906)<br>D-statistic: Women 2.810; Men 2.790; R<sup>2</sup> 0.65 | d graphically | WOMEN: 0.869 (0.865-0.873), MEN: 0.878 (0.875-0.881)<br>D-statistic: Women 2.440; Men 2.470; R<sup>2</sup> 0.59 |  |  |
| LCR009 | Guo<sup>30</sup> | Age; history of chronic respiratory disease; first-degree family history; menopause; benign breast disease | Retrospective | Cox proportional hazards | Chinese female nonsmokers | Lung cancer diagnosis (Oct 2013–Mar 2020) | Assessed graphically | C-statistic: Training 1-yr 0.762; 3-yr 0.718; 5-yr 0.703<br>Validation 1-yr 0.646; 3-yr 0.658; 5-yr 0.650 | – | – | – | – |
| LCR015<sup>31</sup> | Kent & Medway Lung Cancer Risk Tool | Age; activity score; smoking score; any respiratory illness; hypertension; cancer; tuberculosis | Retrospective analysis | Linear regression | Adult residents of Kent & Medway | Primary malignant lung cancer over 6 years | – | Detected 822 vs 581 cases (41.4% increase vs TLHC criteria) | – | – | – | – |
| LCR017 | Chandran<sup>32</sup> | 278 predictors (top: age; smoking; | Retrospective | Machine learning | Adults aged | Lung cancer | Assessed | C-statistic 0.76 (95% CI 0.75– | Assessed | C-statistic Mercy | – | – |
|  |  | race/ethnicity;<br>COPD) |  |  | 45–65 | over 3<br>years<br>(post-<br>index) | graphic<br>ally | 0.78); | graphic<br>ally | EHR 0.81<br>(95% CI<br>0.79–<br>0.84);<br>Optum<br>0.72 (95%<br>CI 0.71–<br>0.74);<br>PPV:<br>Mercy<br>0.97;<br>Optum<br>0.68 |  |  |
| LCR01<br>8 | Ma<sup>33</sup> | Ever-smokers: Age<br>group; area;<br>education; height;<br>BMI; cough;<br>emphysema/chroni<br>c bronchitis; cancer<br>history; ≥2 first-degree relatives<br>with cancer;<br>cigarettes/day;<br>smoking years;<br>inhalation; quit<br>yearsNever-<br>smokers: same<br>minus smoking<br>terms | Prospecti<br>ve | Cox<br>proporti<br>onal<br>hazards | Chines<br>e ever-<br>&<br>never-<br>smoker<br>s | Lung<br>cancer<br>inciden<br>ce | Assesse<br>d<br>graphic<br>ally | 6-year AUC<br>Ever smokers:<br>AUC = 0.778<br>(0.768-0.789)<br>Never<br>smokers: AUC<br>= 0.733 (0.720-0.747) | Assesse<br>d<br>graphic<br>ally | 6-year<br>AUC<br>Ever<br>smokers:<br>AUC =<br>0.774<br>(0.695-0.853)<br>Never<br>smokers:<br>AUC =<br>0.759"<br>(0.675-0.842) | – | – |
| LCR02<br>1 | China<br>NCC-LCm20<br>21<sup>34</sup> | Never-smokers:<br>Age; sex; BMI;<br>family history;<br>chronic respiratory<br>diseases | Prospecti<br>ve | Cox<br>proporti<br>onal<br>hazards | Chines<br>e ever-<br>&<br>never-<br>smoker<br>s | Lung<br>cancer<br>inciden<br>ce up<br>to 3–6<br>years | Slope<br>(3-yr<br>0.975;<br>5-yr<br>0.969;<br>6-yr | AUC (ever):<br>3-yr 0.728;<br>5-yr 0.722;<br>6-yr 0.719;<br>AUC (never): | E/O (3-yr val<br>B):<br>never<br>0.95;<br>ever | AUC<br>(ever):<br>0.752<br>(0.715 -<br>0.789); | 3-yr 0.68<br>(0.51%) | 3-yr<br>0.66<br>(0.51%<br>) |
|  |  | Ever-smokers:<br>Age; BMI;<br>cigarettes/day;<br>years smoked |  |  |  |  | 0.973);<br>E/O 3-yr never<br>0.958;<br>ever<br>0.975 | 0.698 | 1.04 | AUC<br>(never):<br>0.673<br>(0.650 -<br>0.695) |  |  |
| LCR02<br>7 | Yeo <sup>35</sup> | Age; sex; BMI;<br>smoking; alcohol;<br>diabetes; COPD;<br>pulmonary TB;<br>insurance | Retrospec<br>tive | Cox<br>proporti<br>onal<br>hazards | South<br>Korean<br>adults<br>40–90 | Lung<br>cancer<br>over<br>mean<br>8.2<br>years | Assesse<br>d<br>graphic<br>ally | AUC 0.825<br>(95% CI<br>0.810–0.840) | – | – | – | – |
| LCR03<br>1 | LLPv3<br><sup>36</sup> | Age; sex; smoking<br>history; prior<br>pneumonia;<br>asbestos; prior<br>malignancy; family<br>history | Case-Control | Logistic<br>regressi<br>on | UK<br>residen<br>ts 50–<br>75 | Lung<br>cancer<br>within<br>5 years | Slope<br>0.90<br>(95% CI<br>0.75–<br>1.05);<br>CITL<br>LLPv3<br>–0.66;<br>LLPv2<br>–1.31;<br>$\chi^2$<br>improve<br>d | C-statistic 0.81<br>(95% CI 0.79–<br>0.82) | – | – | – | – |
| LCR03<br>4 | ALIGN<br>ED <sup>37</sup> | 17 variables: age;<br>sex; socioeconomic<br>status; smoking<br>status; family<br>history; BMI;<br>BMI×smoking<br>interaction; alcohol<br>misuse; COPD;<br>coronary heart<br>disease; dementia;<br>hypertension; | Combine<br>d data | Logistic<br>regressi<br>on | UK<br>populat<br>ion 55–<br>75 | Lung<br>cancer<br>at 6<br>years | Slope<br>(decile<br>cut-<br>offs) | C-statistic<br>0.804 (95% CI<br>0.799–0.809) | – | C-statistic<br>0.744<br>(95% CI<br>0.723–<br>0.766) | – | – |
|  |  | painful condition; stroke; peripheral vascular disease; previous cancer; previous pneumonia. |  |  |  |  |  |  |  |  |  |  |
| LCR039 | PLCOT-2<sup>38</sup> | Age; Asian ethnicity; education; BMI; COPD; personal history of cancer; family history of (lung) cancer; smoking status; duration of smoking; smoking intensity; quit time (years) | Retrospective | Cox proportional hazard | Taiwan ; multi-center case sources and national datasets | Incident lung cancer within 6 years | PLCOT-2: 1.83 (40–10), 1.46 (30–15), 1.35 (USPST F-21), 1.34 (NELSON) | AUC: 0.755 | | | 66.7% ($\geq 0.0124$) | 67.2% ($\geq 0.0124$) |
| LCR040 | Alexander W Jung<sup>39</sup> | (a) 1,305 binary diagnosis indicators (ICD-10 3rd-level, chapters 1–18, excluding malignant neoplasms); (b) 80 categorical family-history variables (cancer in up to 2nd-degree relatives for each of 20 cancer sites, by relatedness); (c) text-mined basic health factors: alcohol use, smoking, height, weight, blood pressure, and age at first birth | Retrospective | Cox regression | Danish registry 16–86; UK Biobank 50–75 | 1-year lung cancer risk | Assessed graphically | Danish: C-statistic 0.89; Age-sex adjusted 0.67 | Assessed graphically | UKB: C-statistic 0.78 (95% CI 0.77–0.79); Age-sex 0.73 (95% CI 0.72–0.74) | – | – |
| LCR041 | UCL-I<sup>40</sup> | Age; smoking duration; pack-years; BMI; COPD; family history | Combined data | Machine learning | General population | Lung cancer within 5 years | E/O 1.0 (0.92–1.07); Slope ever 1.35 | C-statistic 0.810 (0.800–0.820); Brier ever 0.006 | E/O 1.0 (0.92–1.07) Brier 0.0147 | C-statistic 0.78 (0.77–0.79); | UCL-I 83.9%; UCLFull-I 84.3% | UCL-I 57.7%; UCLFull-I 57.7% |
| LCR046 | Yeh<sup>41</sup> | Age; gender; imaging history (ICD-9-CM codes); medications (ATC groups) | Retrospective | Neural network | Taiwanese population | 1-year lung cancer incidence | Assessed Graphically | AUC (diag+med) 0.902 |  | Testing AUC 0.911 | 0.804 | 0.837 |
| LCR050 | Guo<sup>42</sup> | Age; gender; education; family history; history of tuberculosis; hyperlipidemia | Prospective | Cox proportional hazards | Chinese non-smokers | Lung cancer at 1-, 3-, 5-years | Assessed Graphically | C-statistic: Training 1-yr 0.753; 3-yr 0.752; 5-yr 0.755<br>Validation: 1-yr 0.668; 3-yr 0.678; 5-yr 0.685 | – | – | – | – |
| LCR059 | Survival Stacking Ensemble<sup>43</sup> | Age; years smoking; cigarettes/day; years quit; family history; BMI; sex | Analysis of trial data | Cox regression (stacked ensemble) | Smokers 55–74 yrs (NLST, PLCO) | 3-year lung cancer risk | – | Train 0.789 on PLCO control<br>Test 0.799 on PLCO radiography | – | AUC 0.698 on NLST validation | 33.9% (15%); 76.6% (50%); 92% (75%) | – |
| LCR067 | Boyoung Park<sup>44</sup> | Age (mean-centered<sup>2</sup>); sex; $\sqrt{\text{pack-years}}$; smoking status; | Retrospective | Cox regression | Korean ever-smokers 40–79 | Lung cancer over mean | 1.002 (E/O, training) | Harrell's C-index 0.816 (training) (95% CI: | | | 60.6% (risk >2.1%, train); 60.1% (val) | 83.1% (risk >2.1%, train); |
|  |  | years quit; physical activity; alcohol; BMI categories; COPD; emphysema; pneumoconiosis; IPD |  |  |  | 6.6 years | 0.989 (E/O, validation) | 0.810-0.822)<br>Harrell's C-index 0.816 (validation) (95% CI: 0.806-0.826) |  |  |  | 83.0% (val) |

**AUC:** area under ROC curve; **C-statistic:** concordance statistic; **E/O:** expected/observed ratio; **CITL:** calibration-in-the-large; **PPV:** positive predictive value; **D-statistic:** discrimination measure; **R<sup>2</sup>:** explained variation; **VTE:** venous thromboembolism; **TB:** tuberculosis, **IPD:** interstitial pulmonary disease

These models were developed and validated across diverse populations, including Asian ever-smokers, general populations in the UK and China, female non-smokers. Samples also varied and included clinical trials and health-system registries; prediction methodology involved both traditional regression and machine-learning approaches.

Of the 20 risk models, six followed prospective designs (e.g. Shanghai-LCM, Guo 2021, OWL), ten had retrospective designs (e.g. Levi, CanPredict, Guo 2023), two used combined sets of data (ALIGNED, UCL-I), one used a case-control design (LLP-V3), and one analyzed trial data (Survival Stacking Ensemble).

Regression-based approaches predominated in model development: eleven studies fitted Cox proportional-hazards models (e.g.: Shanghai-LCM, CanPredict, Jung, Park), two used logistic regression (LLPv3, ALIGNED) and one linear regression (Kent & Medway Lung Cancer Risk tool). Machine-learning methods were applied in four studies, ensemble trees (Levi), a mixed ensemble (UCL-I), machine learning (Chandran) and a neural network (Yeh), and one study built stacked Cox ensembles on trial data (Survival Stacking Ensemble).

Optimism-corrected internal values of discrimination were reported for almost all models and spanned from moderate to excellent: the neural-network model by Yeh reached the highest c-statistic (0.902), while and the regression-based CanPredict had similar values (0.897 in women and 0.904 in men); most Cox or logistic models clustered between 0.75 and 0.83, for example Shanghai-LCM 0.78, LLPv3 0.81, UCL-I 0.81, Yeo 0.83 and Park 0.82, while Ma, NCC-LCm and the survival-stacking ensemble lay in the mid-0.70s.

By contrast, quantitative metrics related to internal calibration appeared only in five studies. Calibration slopes were close to unity in Shanghai-LCM (0.98), NCC-LCm (0.97), LLPv3 (0.90) and Park (E/O 1.00); UCL-I reported an E/O ratio of 1.0 but a slope of 1.35, indicating some mis-calibration, whereas Shanghai-LCM’s E/O ratio of 0.70 suggested under-prediction The remaining models stated that calibration was inspected graphically. Overall, discrimination was generally acceptable to strong across models and formal internal calibration assessment was often lacking.

External discrimination metrics were available for ten models. Discrimination (C-statistic/AUC), and were generally lower than corresponding values from internal validation. They ranged from 0.70 for the stacked Cox ensemble tested in NLST to 0.88 for CanPredict in men and 0.87 in women; intermediate values were 0.70 for Shanghai-LCM, 0.67–0.75 for NCC-LCm, 0.74 for ALIGNED, 0.76–0.77 for Ma, 0.78 for UCL-I on PLCO, 0.82 for Park, and 0.72–0.81 for Chandran across two health-care datasets. Quantitative metrics of calibration appeared in only three studies: NCC-LCm reported slopes 0.95–0.97 and E/O ratios 0.95–1.04, UCL-I an E/O ratio of 1.00, and Park an E/O of 0.99; other models offered graphical assessments or none.

Overall, regression-based models achieved moderate to excellent discrimination (C-statistics 0.71–0.90) with generally good internal calibration when evaluated. Machine-learning approaches performed comparably, with some models (Yeh neural network, CanPredict) surpassing regression-based methods in AUC. Nevertheless, external validation was unevenly applied yet usually confirmed reasonably preserved discrimination. ThresholdLbased sensitivity and specificity varied according to chosen cut-points.

Among the 21 screening models that incorporated biomarkers (Table 2), nine used prospective designs, six case-control, two in retrospective, two used pooled data, and one analysis of trial data. Logistic regression was the most common modelling approach (8 models); four used time-to-event Cox-based methods, six studies applied machine-learning algorithms such as XGBoost, Random Forest or multi-algorithm ensembles; and one employed a flexible Royston–Parmar survival model. As expected, logistic regression predominated in case-control and nested case-control designs, whereas machine-learning and survival models were mainly used in prospective designs.

**Table 2.** Risk-based lung cancer screening models incorporating biomarker data.

|  |  |  |  |  |  |  |  |  |  |  |  |  |
| --- | --- | --- | --- | --- | --- | --- | --- | --- | --- | --- | --- | --- |
| Study Number | Risk Prediction Model | Risk Factors | Study Design | Model Type | Targeted Population | Prediction Type | Predictive Performance |  |  |  | Sensitivity (threshold) | Specificity (threshold) |
|  |  |  |  |  |  |  | Internal Validation |  | External Validation |  |  |  |
|  |  |  |  |  |  |  | Calibration | Discrimination | Calibration | Discrimination |  |  |
| LCR012 | Michael K. Gould <sup>45</sup> | Sociodemographic; smoking history; BMI; comorbidities; hospitalizations; CBC; coagulation; electrolytes; hepatic & kidney function; spirometry | Case–control | XGBoost; logistic regression | KPSC members aged 45–90 (NSCLC cases N=6,505; controls N=189,597) | NSCLC diagnosis | Calibration plot (graphical) | AUC: MES 0.856 (95% CI 0.841–0.871); LR 0.840 (0.824–0.858);<br>mPLCom2012: 0.791 (0.771–0.810) | – | – | 40.1% (at 95% specificity) | 95% |
| LCR014 | Fahrman 4MP <sup>46</sup> | PLCom2012 variables + circulating protein biomarkers (4MP) | Case–control | Logistic regression | PLCO ever-smokers aged 55–74 | Lung cancer within 6 yrs (subset 1 yr) | – | AUC 0.85 (95% CI 0.82–0.88) | – | – | 83.5% (≥1.7%) ; 88.4% (≥1.0%) | 69.3% (≥1.7%) ; 56.2% (≥1.0%) |
| LCR020 | Siqi Zhang <sup>47</sup> | Demographics ; lifestyle; health; lab data; work environment; anthropometry | Prospective cohort | Logistic regression; Naive Bayes; XGBoost; RF | UK Biobank participants (n=467,888) | Incident primary invasive lung cancer | Assessed Graphically | Training AUCs: LR 0.839; NB 0.877; XGB 0.998;<br>RF 1.000<br>Test AUCs: LR 0.848; NB 0.879; |  |  | 68% (95% CI 0.63–0.72) | 66% (95% CI 0.66–0.66) |
|  |  |  |  |  |  |  |  | XGB<br>0.998;<br>RF 0.968 |  |  |  |  |
| LCR0<br>25 | MetaPR<br>S<sup>48</sup> | SNP-based<br>PRS +<br>sociodemogra<br>phic &<br>behavioural | Pooled<br>data | Cox<br>regressi<br>on | UKB &<br>PLCO<br>participants<br>without<br>prior cancer | 10–12 yr<br>lung<br>cancer risk | – | C-index:<br>UKB 0.590 | – | C-index:<br>PLCO<br>0.580 | – | – |
| LCR0<br>26 | Cortés-Ibáñez<sup>49</sup> | Age; sex;<br>BMI;<br>smoking;<br>GDF-15; IL-6;<br>CRP; NT-proBNP;<br>spirometry | Analysis<br>of<br>Randomiz<br>ed trial | Logistic<br>regressi<br>on | LUSI trial<br>smokers<br>aged 50–69 | Up to 11.8<br>yr lung<br>cancer risk | – | Model 1:<br>58.2 (44.4–<br>71.4);<br>+GDF-<br>15/IL-6:<br>64.9 (49.5–<br>77.8)<br>Model 2:<br>65.6 (47.6–<br>82.1);<br>+GDF-<br>15/IL-6:<br>73.5 (45.9–<br>87.7) | – | – | – | – |
| LCR0<br>32 | Horsfall<sup>50</sup> | Scenario 1:<br>demographics;<br>smoking;<br>family history;<br>SES;<br>conditions;<br>exposure<br>Scenario 2: +<br>FEV<sub>1</sub>;<br>alcohol; | Prospecti<br>ve | Royston<br>–<br>Parmar<br>PH<br>model | UKB<br>participants<br>≥ 40 yrs<br>without<br>lung cancer | 7-yr<br>incident<br>lung<br>cancer | – | AUCs:<br>S1 0.805<br>(0.794–<br>0.816);<br>S2 0.811<br>(0.800–<br>0.822);<br>S3 0.815 | – | – | – | – |
|  |  | waistScenario 3: + liver tests;<br>urateScenario 4: all above |  |  |  |  |  | (0.804–0.826);<br>S4 0.819 (0.808–0.830) |  |  |  |  |
| LCR037 | HUNT Lung □ S NP model <sup>51</sup> | Clinical (8 vars): sex; age; BMI; pack-yrs; cig/day; quit yrs; indoor smoke; coughGenetic (22 SNPs) | Prospective | Logistic regression | Ever-smokers | 6-yr lung cancer risk | "Average O/E = 0.87 (Discovery) Assessed Graphically" | AUC = 0.875 (95% CI 0.854–0.896) | Average O/E = 0.96 (Validation) | AUC = 0.916 (95% CI 0.880–0.948) | 73.8% (68.0–79.4) Discovery; 76.9% (65.5–87.5) Validation (top 16th percentile) | 84.3% (83.96–84.65) Discovery; 88.8% (88.09–90.08) Validation (top 16th percentile) |
| LCR038 | Anjun Chen <sup>52</sup> | 29 clinical & biomarker vars (e.g. emphysema; inflammation; albumin/globulin; CEA; CRP; NSE; nodules; symptoms) | Retrospective | XGBoost; RF; SVM; KNN | Patients ≥ 30 yrs at Guilin Medical Univ. hospital | Binary lung cancer risk | Assessed Graphically | Internal discrimination – AUROC: XGBoost 0.819;<br>RF 0.826; SVM 0.813;<br>KNN 0.760 | – | – | 80% (threshold not specified) | – |
| LCR045 | LunaCAM-S <sup>53</sup> | ctDNA methylation markers (7-marker panel: SHOX2, HOX | Case–control | Logistic regression | Lung cancer, benign pulmonary disease, | Early-stage lung cancer detection & | – | Training AUC 0.913 (0.900–0.931) | – | Validation AUC 0.901 (0.875–0.937) | 90.6% (88.2–94.1) | 61.9% (56.0–68.0) |
|  |  | genes, etc.) |  |  | healthy controls in China | classification |  |  |  |  |  |  |
| LCR049 | Haixin Yu<sup>54</sup> | Genetic risk score (31 SNPs); methylation score (151 CpGs); pack-yrs | Nested case-control | Logistic regression | ESTHER cohort aged 50–75 (Germany) | 14-yr incident lung cancer | – | AUC: All 0.810 (0.764–0.857);<br>Ever 0.737 (0.681–0.792);<br>Heavy 0.654 (0.576–0.732) | – | – | – | – |
| LCR054 | iPRS<sup>55</sup> | 299 SNPs ($p < 5 \times 10^{-8}$ main/interaction); pack-yrs | Retrospective trial | Logistic regression | PLCO participants with genotype & smoking history | Binary lung cancer occurrence | Assessed Graphically: Brier: Model 1 iPRS 6.2 [5.6–6.9]; Model 2 PRS slope 6.3 [5.5–7.0] | AUC: Model 1 iPRS 0.85; Model 2 PRS 0.83 | – | – | – | – |
| LCR055 | TNSF-SQ<sup>56</sup> | Age; BMI; COPD; education; family history; 9 SNPs (TNSF-SQ)TNSF-SQNG: same without SNPs | Case-control | Logistic regression | Never-smoking females 55–70 (dev); 55–74 (val) in Taiwan | 6-yr lung cancer risk | – | Training AUC: 0.770 (95% CI 0.749–0.791) | – | Test AUC: 0.714 (95% CI 0.660–0.768) | TNSF-SQ: Sens 29.7%; 27.0%; 15.3% (p=0.0134–0.0200) | TNSF-SQ: Spec 94.7%; 96.1%; 98.2% (p=0.0134–0.0200) |
| LCR0<br>57 | Hung<sup>57</sup> | Age; sex;<br>race;<br>education;<br>BMI;<br>smoking;<br>COPD;<br>personal &<br>family history;<br>PRS (128<br>variants) | Pooled<br>data<br>analysis | Logistic<br>regression | European-<br>ancestry<br>screening-<br>eligible<br>cohorts | 5-yr &<br>lifetime<br>lung<br>cancer<br>absolute<br>risk | Assessed<br>Graphically | AUC 0.832<br>(95% CI<br>0.811–<br>0.853)<br>Ever<br>smokers:<br>RF + PRS<br>0.786<br>(0.762–0.809)<br>Non-smokers RF +<br>PRS: 0.687<br>(0.628–0.746) | - | – | – | – |
| LCR0<br>58 | INS<sup>58</sup> | INS proteins<br>(e.g. CASP8,<br>CCL11,<br>CDCP1...);<br>INS-pack-yrs<br>adds pack-years | Prospective cohort | Machine<br>learning | Saarland<br>ever-smokers<br>aged 50–75<br>(ESTHER) | Incident<br>lung<br>cancer risk | – | Training<br>AUC:<br>INS 0.771<br>(0.713–<br>0.828);<br>Validation<br>AUC:<br>INS 0.742<br>(0.667–<br>0.818);<br>Training<br>INS-pack-yrs 0.811<br>(0.760–<br>0.863) | – | - | – | – |
|  |  |  |  |  |  |  |  | Validation<br>INS-pack-yrs 0.782<br>(0.711–<br>0.854) |  |  |  |  |
| LCR0<br>60 | MTL-Cox <sup>59</sup> | Smoking<br>status; age;<br>cig/day; past<br>smoking;<br>NEUT1; fruit<br>intake; RBC1;<br>PCT1; Cyc1;<br>IRF1;<br>HLSRC1;<br>ApoA1;<br>LDL1;<br>BASOP1;<br>BASO1 | Prospecti<br>ve | Multi-task<br>Cox<br>model | UKB<br>participants<br>aged 37–73 | 1-, 3-, 5-,<br>7-yr lung<br>cancer risk | – | AUC:<br>MTL-Cox<br>0.822;<br>Cox 0.821;<br>Weibull<br>0.747;<br>Cox-Lasso<br>0.821 | – | – | Sens:<br>MTL-Cox<br>0.696;<br>Cox<br>0.752;<br>Weibull<br>0.659;<br>Cox-Lasso<br>0.752 | Spec:<br>MTL-Cox<br>0.802;<br>Cox<br>0.750;<br>Weibull<br>0.721;<br>Cox-Lasso<br>0.750 |
| LCR0<br>62 | Jia <sup>60</sup> | PRS (19<br>SNPs); +<br>family history | Prospecti<br>ve | Cox<br>regressi<br>on | UKB<br>Europeans | 5.8 yr lung<br>cancer<br>incidence | – | - | – | – | – | – |
| LCR0<br>63 | Jacobsen<br><sup>61</sup> | Age; sex;<br>education;<br>smoking;<br>pack-yrs;<br>AHRR<br>(cg05575921) | Prospecti<br>ve | Logistic<br>regressi<br>on | Copenhagen<br>Heart Study<br>smokers | 10-yr lung<br>cancer<br>diagnosis | Hosmer–<br>Lemesho<br>w test | – | – | AUC 0.70<br>(95% CI<br>0.69–0.72) | 0.74<br>(95% CI<br>0.63–<br>0.83) | 0.70<br>(95% CI<br>0.68–<br>0.71) |
| LCR0<br>66 | Lian <sup>62</sup> | Demographics<br>; lifestyle;<br>clinical; NMR<br>biomarkers<br>(e.g., HbA1c;<br>MUFA;<br>GlycA;<br>omega-3; | Prospecti<br>ve | Machin<br>e<br>learning | UKB with<br>plasma<br>NMR data | 11.3 yr<br>incident<br>lung<br>cancer | – | AUC:<br>0.702<br>(0.691 -<br>0.713) | – | – | – | – |
|  |  | amino acids; lipoproteins; metabolites) |  |  |  |  |  |  |  |  |  |  |
| LCR068 | Wei CKB<sup>63</sup> | PRS-21; age; education; BMI; COPD; personal & family cancer history; ancestry PCs | Prospective | Cox regression | CKB never-smoking Chinese women 30–79 | 10.53 yr incident lung cancer | – | AUC: combined 0.711;<br>Conventional 0.697;<br>PRS alone 0.576 | – | – | – | – |
| LCR069 | Yao<sup>64</sup> | 28 metabolites (e.g. pregnenolone sulfate; lactic acid; kynurenine; serotonin; DHA; acylcarnitines; etc.) | case-control | LASSO logistic regression | Chinese centers: controls; benign nodules; stage I adenocarcinoma | Benign vs malignant nodule differentiation | – | AUC: Discovery 0.933;<br>Internal 0.915 | – | AUC: External 0.945 | 86.8% Discovery;<br>86.7% Internal;<br>81.0% External | 85.9% Discovery;<br>81.1% Internal;<br>97.9% External |
| LCR073 | Wang<sup>65</sup> | Plasma CTA-384D8.35; Plasma PGM5-AS1; Exosome CTA-384D8.35; Exosome PGM5-AS1; Exosome SFTA1P; log10-CEA; log10-CA125; SCC; NSE | case-control | logistic regression | Adults with NSCLC (LUAD/LUSC) vs healthy controls in Chinese hospitals | Diagnosis of NSCLC | Assessed graphically | training (AUC=0.972)<br>validation (AUC=0.973) | - | - | - | - |

Internal discrimination varied by approach. MachineLlearning models frequently achieved the highest values, Siqi Zhang’s XGBoost and Random Forest reported training AUCs of 0.99–1.000, though overfitting remains a concern. RegressionLbased models typically achieved AUCs or C-statistics between 0.80 and 0.86 (e.g: Gould’s MES AUC 0.86; Fahrmann’s four-protein panel AUC 0.85). Polygenic risk scores alone performed modestly (AUCs ∼0.58–0.62) but improved when combined with clinical variables (e.g., Wei CKB model AUC 0.71). Calibration metrics were less consistently reported: Nguyen et al. (HUNT lung – SNP Model) reported average O/E ratios of 0.87 (discovery) and 0.96 (validation), whereas most others provided only graphical calibration plots or Hosmer–Lemeshow tests.

External validation was reported for six models, and again were lower than internal validation values (MetaPRS, Hunt Lung SNP, LunaCAM-S, TNSF-SQ, Jacobsen et al., and Yao et al.). Discrimination varied from a C-statistics of 0.58 for the MetaPRS score to an AUC 0.916 for the HUNT Lung-SNP model, Yao metabolomic panel AUC 0.945, LunaCAM-S AUC 0.901, TNSF-SQ AUC 0.714, and Jacobsen AUC 0.74. Two models exceeded an AUC of 0.90 (HUNT Lung-SNP, Yao et al., LunaCAM-S). Calibration was seldom quantifiedin external validation; Nguyen reported an O/E ratio of 0.96. Heterogeneity in datasets, metrics, and reporting limits direct comparison across models.

Overall, Internal discrimination covered a broad span, from an AUC 0.591 for a 19-variant polygenic score to 1.00 for a random-forest model in UK Biobank, with most models lying between 0.80 and 0.88. For the six models that reported external validation, C-index/AUC values ranged from 0.58 (MetaPRS) to 0.95 (metabolomic panel by Yao), and three exceeded 0.90. Calibration was rarely quantified: Nguyen reported an observed-to-expected ratio of 0.96, while most other studies provided only graphical checks. Reported operating points showed wide variation, sensitivity spanned 15.3–29.7% for TNSF-SQ (specificity ≥ 94.7 %) to 90.6 % for a ctDNA methylation panel (specificity 61.9 %), with the metabolomic model achieving 81.0 % sensitivity at 97.9 % specificity.

***Table 3*** reports on the 27 models in that predicted malignancy risk in screen-detected pulmonary nodules. Among these, analyses of lung-cancer screening trials dominated, with fourteen having analyzed trial data, namely LCi; Brock & LCP-CNN; S-RRL/B-RRL; Lu; qADD++; Rundo; Sybil; Venkadesh 1; Venkadesh 2; Kaiwen Xu; Trajanovski; Ebrahimpour; Paul; and CXR-LC. Seven models stem used retrospective designs (Huijie, Kaeum, Chang, Yao, cLung-RADS, Jieke Liu and Gao). Two had combined data from multiple sources (Dahai Liu and INTEGRAL-Radiomics), and three used prospective designs (LCP-CNN, Xiao and Wu).

**Table 3.** Malignancy risk prediction models for screen-detected pulmonary nodules.

|  |  |  |  |  |  |  |  |  |  |  |  |  |
| --- | --- | --- | --- | --- | --- | --- | --- | --- | --- | --- | --- | --- |
| Study Number | Risk Prediction Model | Risk Factors | Study Design | Model Type | Targeted Population | Prediction Type | Predictive Performance |  |  |  | Sensitivity (threshold) | Specificity (threshold) |
|  |  |  |  |  |  |  | Internal Validation |  | External Validation |  |  |  |
|  |  |  |  |  |  |  | Calibration | Discrimination | Calibration | Discrimination |  |  |
| LCR 006 | LCi(final)<sup>66</sup> | Sex; BMI; smoking intensity; duration; quit time; family history; emphysema score; mean lung density; bronchial wall thickness; aorta calcium volume & density; nodule longest & perpendicular diameters; upper-lobe location; spiculation | Analysis of trial data | Cox regression | Smokers | Lung cancer incidence (1–5 year) | Slope (NLST):<br>1-yr 0.804;<br>2-yr 0.650;<br>3-yr 0.609;<br>5-yr 0.603 | AUC (NLST):<br>1-yr: 0.937 (0.920–0.954)<br>3-yr: 0.859 (0.843–0.875)<br>5-yr: 0.840 (0.826–0.855) | Slope (MILD):<br>1-yr: 1.240;<br>2-yr 0.972;<br>3-yr 0.890;<br>5-yr 0.854 | AUC (MILD):<br>1-yr: 0.902 (0.810–0.995)<br>3-yr: 0.808 (0.737–0.879)<br>5-yr: 0.799 (0.739–0.858) | 1-yr 83.9%;<br>2-yr 70.3%;<br>3-yr 59.4%;<br>5-yr 51.7% (90% quantile) | 1-yr 90.9%;<br>2-yr 91.2%;<br>3-yr 91.5%;<br>5-yr 91.8% (90% quantile) |
| LCR 008 | Brock & LCP-CNN<sup>67</sup> | CT imaging data (CNN); age; sex; family history; emphysema; nodule size; spiculation; count; location; part-solid | Analysis of trial data | Logistic regression (Brock); CNN (LCP-CNN) | Ever-smokers aged 55–74 | Nodule malignancy risk | – | AUC: LCP-CNN 0.936 (0.926, 0.945); | – | – | – | – |
| LCR 010 | Huijie <sup>68</sup> | Age group; sex; smoking & drinking history; symptoms; CT-based pixel intensity & texture | Retrospective | Logistic regression; CNN | Chinese general population | Malignant vs benign nodules | – | AUC: Clinical 0.823 (0.799 - 0.847)<br>(train); 0.701 (0.668-0.733) (test); CNN 0.984 (0.971-0.986) (test) | – | – | Clinical : 87.3% (train); 80.2% (test) CNN: 96.1% (no CV); 94.7% (5-fold CV) | Clinical: 67.7% (train); 61.8% (test) CNN: 96.0% (no CV); 93.3% (5-fold CV) |
| LCR 011 | S-RRL / B-RRL <sup>69</sup> | Deep radiomics (32 ResNet-18 features); age; sex; family history; emphysema | Analysis of trial data | Radiomics-based RL | Ever-smokers | Malignant vs benign nodules (≤7 yr follow-up) | – | AUC: S-RRL 0.88 (0.85, 0.91);<br>B-RRL 0.86 (0.83, 0.90);<br>S-RRL at diagnosis 0.90 (0.87, 0.93);<br>D-RRL 0.89 (0.86, 0.92) | – | – | – | – |
| LCR 013 | CXR-LC <sup>70</sup> | Chest radiograph; age; sex; | Analysis of trial data | CNN | Age 55–74 general population | Lung cancer incidence | E/O 1.01 (95% CI 0.87–1.14) | CXR-LC 12y: 0.755 (0.723– | – | CXR-LC: 0.659 (0.622– | 71.4% (6-yr, NLST | 62.7% (6-yr) |
|  |  | current<br>smoking |  |  |  | (6- & 12-<br>yr) |  | 0.786);<br>CXR-LC<br>6y: 0.728<br>(0.682–<br>0.774);<br>CXR-LC<br>+ CMS<br>12y: 0.759<br>(0.729–<br>0.790);<br>CXR-LC<br>+ CMS<br>6y: 0.738<br>(0.695–<br>0.781);<br>CXR-LC<br>+<br>PLCOm20<br>12 12y:<br>0.790<br>(0.763–<br>0.817);<br>CXR-LC<br>+<br>PLCOm20<br>12 6y:<br>0.767<br>(0.728–<br>0.806) |  | 0.695);<br>CXR-LC<br>+<br>PLCOm2<br>012 6y:<br>0.686<br>(0.651–<br>0.721) | eligible<br>) |  |
| LCR<br>016 | Kaeum<sup>71</sup> | Age; sex;<br>BMI; smoking | Retrospe<br>ctive | Logistic<br>regressi | South Korean<br>general | Cancer<br>within 5 yr | Brier:<br>clinical | AUC:<br>Clinical | – | – | – | – |
|  |  | history; SUV metrics; MH index |  | on | population |  | 0.119; SUVR 0.131; MH 0.125; Clinical+S UVR 0.100; clinical+MH 0.100 | 0.798 (0.725-0.872);<br>SUVR 0.681(0.587-0.775);<br>MH 0.825 (0.763-0.887);<br>Clinical+S UVR 0.867 (0.802-0.931); Clinical+MH 0.901 (0.848-0.954) |  |  |  |  |
| LCR 019 | Chang <sup>72</sup> | Demographics (e.g., diabetes, COPD); WBC; BUN; age; thyroid; liver; digestive; LDCT features (spiculation; part-solid; GGO; count; solidity; calcification) | Retrospective | Regularized regression | Taiwanese general population | Stage I lung cancer diagnosis | – | AUC 0.93 ($\lambda=0.037$, $\alpha=1$) | – | – | 85% ($\lambda=0.037$, $\alpha=1$) | 100% ($\lambda=0.037$, $\alpha=1$) |
| LCR | Venkadesh | Raw CT | Analysis | 2D & | Ever-smokers | Nodule | – | AUC 0.91 | – | AUC 0.93 | 71% | 90% |
| 022 | 1<sup>73</sup> | voxels | of trial data | 3D CNN |  | malignancy risk |  | (95% CI 0.90–0.92) |  | (95% CI 0.89–0.96) | (internal); 84% (external) | (internal & external) |
| LCR 023 | INTEGRAL-Radiomics<sup>74</sup> | Age; years smoked; sex; FHLC; cigarettes/day; BMI; former smoking; COPD; 134 radiomic features | Combined data | XGBoost; LASSO; RF | Ever-smokers | Malignant vs benign nodules | E/O 1.02; CITL 0.69 | XGBoost: 0.933 (95% CI: 0.923–0.944)<br>LASSO: 0.930 (95% CI: 0.914–0.946)<br>Random Forest: 0.916 (95% CI: 0.904–0.929) | – | – | ≥10% risk: 74.8% | ≥10% risk: 94.7% |
| LCR 028 | Yao<sup>75</sup> | Age; sex; smoking; STK1p; AFP; CEA; BMI; hypertension; hyperglycemia; dyslipidemia; nodule size; type; count | Retrospective | Logistic regression | Chinese general population | Cancer incidence within 3 yr | E/O 1.00; CITL 0.00; slope 1.00 | AUC 0.91 (95% CI 0.86–0.96) | – | – | 84.6% | 92.3% |
| LCR 029 | DL-based cLung-RADS<sup>76</sup> | CT imaging; cLung-RADS category; DL risk scores | Retrospective | 3D-DCNN | Chinese general population | GGN malignancy risk | – | AUC: Train 0.753 (0.526– | – | – | – | – |
|  |  |  |  |  |  |  |  | 0.980);<br>Validation<br>0.734<br>(0.585-0.884) |  |  |  |  |
| LCR<br>030 | Dahai Liu<sup>77</sup> | Epidemiologic<br>al data; CNN<br>score | Combine<br>d trial<br>analysis | Regress<br>ion;<br>CNN | Patients with<br><30 mm<br>nodules on<br>LDCT | Malignant<br>vs benign<br>nodules | Assessed<br>Graphicall<br>y | Training<br>cohort:<br>91.6%<br>(95% CI:<br>89.4%–<br>93.5%) | Assesse<br>d<br>Graphic<br>ally | External<br>validation<br>cohort:<br>88.3%<br>(95% CI:<br>83.1%–<br>92.3%) | 82.3% | 84.6% |
| LCR<br>033 | LCP-CNN<sup>78</sup> | CT imaging<br>data | Prospecti<br>ve | CNN | General<br>population | Benign vs<br>malignant<br>nodules | – | AUC<br>(NLST):<br>92.1%<br>(95% CI<br>91.2–92.9) | – | AUC:<br>VUMC<br>83.5%<br>(75.4-90.7%);<br>OUH<br>91.9%<br>(88.7-94.7%) | – | – |
| LCR<br>035 | Lu<sup>79</sup> | 20 semantic<br>CT features<br>(e.g. shape;<br>texture;<br>spiculation;<br>attachment;<br>fibrosis;<br>emphysema) | Analysis<br>of trial<br>data | Logistic<br>regressi<br>on | Ever-smokers<br>aged 55–74 | Malignant<br>vs benign<br>nodules | – | AUC:<br>T0 0.871<br>(0.809-0.933); T1<br>0.917<br>(0.869-0.966); T2<br>0.941<br>(0.899-0.983) | – | – | T0<br>77.5%;<br>T1<br>77.3%;<br>T2<br>82.0% | T0<br>82.8%;<br>T1<br>89.4%;<br>T2<br>97.8% |
| LCR<br>036 | qADD++<sup>80</sup> | Demographics<br>; radiological<br>descriptors; 11<br>radiomic features | Analysis<br>of trial<br>data | SVM<br>(linear) | Ever-smokers<br>aged 55–74 | Malignant<br>vs pre-<br>/benign<br>nodules | – | AUC:<br>invasive<br>vs<br>preinvasiv e 0.901<br>±0.001,;<br>Invasive<br>vs benign<br>0.909<br>±0.001;<br>Invasive<br>vs<br>all0.877±0<br>.001 | – | – | – | – |
| LCR<br>043 | Rundo <sup>81</sup> | 2D shape;<br>GLCM<br>(correlation,<br>cluster shade);<br>GLSZM (size-<br>zone non-<br>uniformity) | Analysis<br>of trial<br>data | Logistic<br>regressi<br>on | High-risk<br>smokers | Malignant<br>vs benign<br>nodules | – | AUC:<br>radiomic<br>0.747 ±<br>0.015;<br>Blinded<br>0.564;<br>SMOTE<br>0.817 ±<br>0.003;<br>Blinded<br>0.556 ±<br>0.007 | – | – | Blinded<br>0.102;<br>SMOT<br>E<br>0.670;<br>blinded<br>set<br>0.462 | Blinde<br>d<br>0.978;<br>SMOT<br>E<br>0.862;<br>blinded<br>0.623 |
| LCR<br>044 | Sybil <sup>82</sup> | LDCT<br>features | Analysis<br>of trial<br>data | CNN | NLST<br>smokers;<br>MGH &<br>CGMH<br>screening<br>cohorts | Lung<br>cancer risk<br>(6 yr) | – | NLST test<br>set: 0.75<br>(95% CI,<br>0.72-0.78) | – | MGH:<br>0.81<br>(95% CI,<br>0.77-<br>0.85).<br>CGMH:<br>0.80<br>(95% CI,<br>0.75- | – | – |
|  |  |  |  |  |  |  |  |  |  | 0.86). |  |  |
| LCR<br>048 | Venkadesh<br>2<sup>83</sup> | 3D CT blocks<br>(current ±<br>prior);<br>volumetric<br>segmentation;<br>time between<br>scans | Analysis<br>of trial<br>data | 3D-CNN | General<br>population | 3-yr<br>malignancy<br>risk | – | AUC:<br>current+pr<br>ior 0.98<br>(0.97-0.98);<br>Latest<br>0.95 (0.94-0.96) | – | AUC<br>(DLCST):<br>0.97<br>(0.95-1.00) vs<br>0.96<br>(0.93-0.99) ;<br>MILD:<br>0.99<br>(0.98-1.00_ vs<br>0.98<br>(0.96-0.99);<br>benign<br>subsets:<br>0.94<br>(0.89-0.98) vs<br>0.89<br>(0.84-0.95) | – | – |
| LCR<br>051 | Jieke Liu<br>84 | CT features;<br>age | Retrospe<br>ctive | Logistic<br>regressi<br>on | Cancer<br>Hospital in<br>China | Adenocarci<br>noma vs<br>benign<br>nodules | H-L test:<br>P=0.650<br>(train);<br>0.998 (val) | AUC:<br>radiomic<br>0.915<br>(0.859-<br>0.972);<br>nomogram<br>0.919<br>(0.861 – | – | AUC:<br>radiomic<br>0.976<br>(0.944 –<br>1.00);<br>nomogra<br>m 0.976<br>(0.941 – | 92% | 85.4% |
|  |  |  |  |  |  |  |  | 0.977)<br>(validation<br>) |  | 1.00)<br>(validation<br>n) |  |  |
| LCR<br>052 | Ebrahimpo<br>ur<sup>85</sup> | Radiomic<br>(intensity,<br>shape, texture,<br>filter, delta<br>T1–T0) | Analysis<br>of trial<br>data | Gradien<br>t<br>Boostin<br>g<br>(MultiS<br>urf) | NLST positive<br>LDCT<br>participants | Cancer risk<br>(1–2 yr) | – | AUC:<br>train 0.85<br>± 0.03 | – | AUC:<br>validation<br>0.76 | – | – |
| LCR<br>056 | Gao<sup>86</sup> | Deep learning<br>CT features;<br>age;<br>education;<br>BMI; cancer<br>history; family<br>history;<br>tobacco status;<br>quit time;<br>pack-years | Retrospe<br>ctive | Neural<br>network | NLST &<br>VLSP<br>participants | Cancer risk<br>within 2 yr | – | AUC:<br>train 0.88<br>± 0.02 | – | AUC:<br>validation<br>0.91<br>(95% CI<br>0.85–<br>0.95) | – | – |
| LCR<br>061 | Xiao<sup>87</sup> | Cooking<br>fumes;<br>depression;<br>hypertension;<br>nodule<br>diameter;<br>density;<br>margin | Prospecti<br>ve | Cox<br>regressi<br>on | Chinese adults<br>40–74<br>undergoing<br>LDCT | Cancer<br>incidence<br>over 6.1 yr | – | Harrell's<br>C 0.847<br>(95% CI<br>0.790–<br>0.904) | – | Harrell's<br>C 0.814<br>(95% CI<br>0.759–<br>0.869) | – | – |
| LCR<br>064 | Wu<sup>88</sup> | Age; mean<br>diameter;<br>calcification;<br>pleural<br>involvement;<br>edge; density;<br>interactions<br>(age×density;<br>age×diameter; | Prospecti<br>ve | Logistic<br>regressi<br>on | High-risk<br>Chinese adults | Cancer<br>within 6<br>mo of<br>LDCT | Assessed<br>Graphicall<br>y | 0.868<br>(95% CI<br>0.839–<br>0.894) | Not<br>reporte<br>d | 0.751<br>(95% CI<br>0.727–<br>0.774) | 85.2%<br>(train<br>3.9%<br>thres);<br>70.5%<br>(val<br>3.1%) | 75.2%<br>(train<br>3.9%);<br>71.0%<br>(val<br>3.1%) |
|  |  | edge×diameter<br>) |  |  |  |  |  |  |  |  |  |  |
| LCR<br>070 | Kaiwen<br>Xu<sup>89</sup> | SAT index &<br>attenuation;<br>SM index &<br>attenuation;<br>age; BMI;<br>emphysema;<br>PLCOM2012<br>factors | Analysis<br>of trial<br>data | Cox<br>regressi<br>on | NLST<br>participants | Lung<br>cancer over<br>median 6.6<br>years | – | Harrell's<br>C<br>Males:<br>0.71 (95%<br>CI 0.69–<br>0.73)<br>Females:<br>0.71 (95%<br>CI 0.68–<br>0.73) | – | – | – | – |
| LCR<br>071 | Trajanovsk<br>i<sup>90</sup> | 2D CT<br>projections;<br>location;<br>radius;<br>confidence;<br>sphericity<br>(Nodule<br>Metadata) | Analysis<br>of trial<br>data | Neural<br>network | NLST, Lahey,<br>Data Science<br>Bowl, UCM<br>scans | Binary<br>malignancy<br>risk | - | - | - | AUC N-Net<br>ensemble:<br>Panel A<br>(Kaggle<br>stage 2):<br>0.886<br>Panel B<br>(LHMC):<br>0.947<br>Panel C<br>(UCM<br>full):<br>0.856<br>Panel D<br>(UCM<br>Radiologi<br>sts):<br>0.875 | At 93%<br>sens:<br>81%<br>spec vs<br>69%<br>(PanCa<br>n); at<br>100%<br>sens:<br>lower<br>spec vs<br>radiolo<br>gists | At<br>80%<br>spec:<br>93%<br>sens vs<br>79%<br>(PanCa<br>n) |
|  |  |  |  |  |  |  |  |  |  | Panel E<br>(Data<br>combined<br>): 0.935 |  |  |
| LCR<br>072 | Paul <sup>91</sup> | Radiomics;<br>nodule size<br>($<6$ mm, $6-16$<br>mm, $\geq 16$<br>mm); sex;<br>smoking;<br>family history | Analysis<br>of trial<br>data | Neural<br>network | NLST LDCT<br>arm nodules | Malignanc<br>y<br>classificati<br>on (1–2 yr) | – | AUC:<br>CNN1<br>0.90;<br>CNN2<br>0.88;<br>CNN3<br>0.90 | – | – | – | – |
| LCR<br>074 | LCRAT+C<br>Tpos <sup>92</sup> | Longest<br>nodule<br>diameter;<br>number of<br>nodules (log);<br>ground-glass<br>nodules;<br>micronodules;<br>nodule<br>location<br>(upper lobes;<br>right middle<br>lobe/lingula);<br>indeterminate<br>margins;<br>interval<br>growth; new<br>nodules;<br>spiculation<br>(strong<br>prevalence/inc<br>idence<br>interaction);<br>plus sex (female) and BMI (overweight/obesity). | Analysis<br>of trial<br>data | Log-<br>binomia<br>l<br>regressi<br>on | Screen-<br>eligible ever-smokers<br>with abnormal<br>LDCT results<br>(NLST<br>criteria). | Immediate<br>lung cancer<br>detection<br>risk at the<br>abnormal<br>screen. | 649<br>observed<br>vs 650<br>predicted<br>($p>0.99$). | AUC 0.86 | - | - | - | - |
| E/O: expected/observed ratio; CITL: calibration-in-the-large; GGO: ground-glass opacity; SUV: standardized uptake value; SUVR: SUV ratio; MH: metabolic heterogeneity index; FHLC: family history of lung cancer; LDCT: low-dose CT; CNN: convolutional neural network; RF: random forest; SVM: support vector machine; XGBoost, LASSO, SMOTE, DCNN, RL: reinforcement learning; NLST, PLCO, CMS, LHMC, MGH, CGMH: study or center acronyms. |  |  |  |  |  |  |  |  |  |  |  |  |

Analytically, conventional Cox or (regularised) logistic regression were used in 10 models; LCi, Kaeum, Chang, Yao, Lu, Rundo, Jieke Liu, Xiao, Wu and Kaiwen Xu. 16 studies used machine-learning approaches, nine of them convolutional neural networks (Brock/LCP-CNN, the Huijie-CNN, CXR-LC, Venkadesh 1, DL-based cLung-RADS, Dahai Liu, the prospective LCP-CNN cohort, Sybil and Venkadesh 2). Seven machine-learning models relied on reinforcement learning, gradient boosting, support-vector machines or ensemble trees: S-RRL/B-RRL (radiomics RL), INTEGRAL-Radiomics (XGBoost/Random Forest), qADD++ (linear SVM), Ebrahimpour (gradient boosting), Gao (feed-forward NN), Trajanovski (projection NN) and Paul (radiomic/tabular NN).

Internal discrimination across models ranged from moderate to high: the LCi Cox model achieved AUCs of 93.7% at one year and 84.0% at five years, whereas the CXR-LC CNN yielded AUCs of 0.73 for six-year incidence and 0.76 for twelve-year incidence. Calibration also varied: the LCi model exhibited calibration slopes below unity (1-year slope = 0.80), suggesting slight risk overestimation, while the CXR-LC model’s O/E ratio was 1.01 (95% CI 0.87–1.14). The INTEGRAL-Radiomics model showed an E/O ratio of 1.02 with a calibration-in-the-large (CITL) of 0.69, alongside a cross-validated AUC of 0.93. In contrast, the Yao logistic model demonstrated perfect internal calibration (E/O = 1.00, CITL = 0.00, calibration slope = 1.00) together with an AUC of 0.91. Overall, several models combined strong discrimination (AUCs ≥ 0.91) with acceptable calibration, though calibration slopes and E/O ratios indicate varying degrees of risk estimation accuracy.

External discrimination AUCs varied across models: the LCi model achieved 0.90 at 1-year, 0.81 at 3-years and 0.80 at 5-years; the CXR-LC model showed 6-year incidence AUCs of 0.66; Venkadesh1 yielded 0.93; Dahai Liu reported 0.88; LCP-CNN reached 0.835 at VUMC and 0.92 at OUH; Sybil recorded 0.81 at MGH and 0.80 at CGMH; Venkadesh2 achieved 0.97 using current + prior CT and 0.89 for the latest CT; Jieke Liu’s radiomic signature and nomogram both gave 0.98; Ebrahimpour obtained 0.76; Gao reported 0.91; Xiao 0.81; and Wu 0.75.

In aggregate, models incorporating quantitative imaging, whether via machine learning or deep-learning feature extraction, achieved very good to excellent discrimination, frequently surpassing AUC of 0.90 internally and maintaining AUC ≥ 0.80 upon external testing. Regression-based models based on large trial data also demonstrated robust discrimination (AUC ∼0.85–0.90) with reliable calibration metrics. Reported sensitivities (61–96%) and specificities (61–98%) reflected differing clinical priorities. The typical decline in discrimination and calibration on external validation underscores the problem of model fitting and the need for local recalibration. The relative stability of the performance of deep-learning algorithms across diverse cohorts is promising for broader clinical deployment, contingent upon rigorous validation in real-world settings.

### Meta-regression analysis of discriminatory performance

***Table 4*** presents the results of meta-regression of discriminatory performance for pre-screening models (24 clusters; 81 AUC values). Larger sample sizes were associated with slightly higher AUC (p = .030). Machine-learning models showed a non-significant increase of 0.037 in AUC compared to regression models (p = 0.307); external validation was associated with a non-significant change of –0.012 (p = 0.439), and inclusion of biomarkers conferred a non-significant difference of +0.033 (p = 0.337). Regression estimates are presented in supplementary Table S2.

**Table 4:** Characteristics of studies at the pre-screening stage (including meta-regression P-values)

|  |  |  |
| --- | --- | --- |
|  | Overall<br>(N=81) | P-value |
| **Sample_Size** |  |  |
| Median [Min, Max] | 70600 [160, 4140000] | 0.0297 |
| **Model_Category** |  |  |
| Regression | 46 (56.8%) | Reference |
| ML | 31 (38.3%) | 0.3071 |
| Neural Network | 0 (0%) | – |
| Other | 4 (4.9%) | 0.8711 |
| **Validation_Type** |  |  |
| internal | 44 (54.3%) | Reference |
| external | 37 (45.7%) | 0.4389 |
| **Biomarkers** |  |  |
| No | 47 (58.0%) | Reference |
| Yes | 34 (42.0%) | 0.3369 |

***Table 5*** presents the post–screening meta-regression results (19 clusters; 62 effect sizes). Here, the association between c-statistic and sample size was not statistically significant (p = 0.759). We observed the general trends in the direction of other associations, but none reached statistical significance: machine-learning (other than neural networks) v. regression (β = +0.035; p = 0.476), neural networks v. regression (β = +0.047; p = 0.415), external v. internal validation (β = –0.001; p = 0.957). Regression estimates are presented in supplementary Table S3.

**Table 5:**
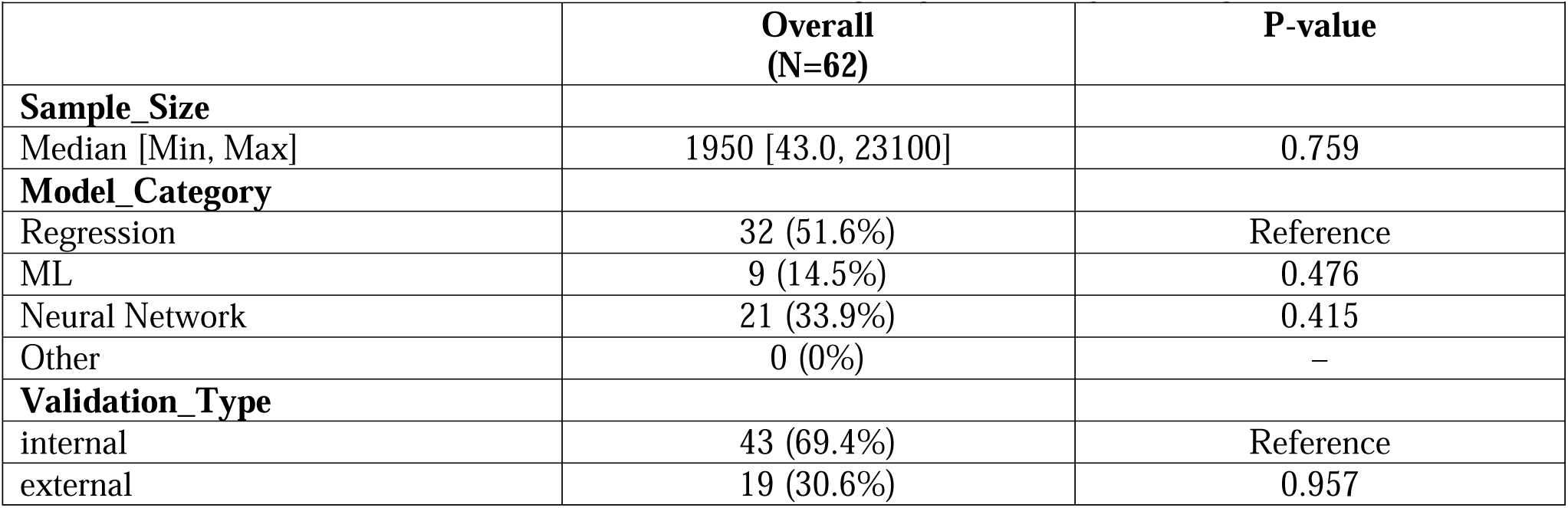
Characteristics of studies at the post-screening stage (including meta-regression P-values)

|  |  |  |
| --- | --- | --- |
|  | **Overall**<br>**(N=62)** | **P-value** |
| **Sample_Size** |  |  |
| Median [Min, Max] | 1950 [43.0, 23100] | 0.759 |
| **Model_Category** |  |  |
| Regression | 32 (51.6%) | Reference |
| ML | 9 (14.5%) | 0.476 |
| Neural Network | 21 (33.9%) | 0.415 |
| Other | 0 (0%) | – |
| **Validation_Type** |  |  |
| internal | 43 (69.4%) | Reference |
| external | 19 (30.6%) | 0.957 |

## Discussion

This systematic review identified 68 new risk prediction models for lung cancer screening developed since 2020. Across all categories, discrimination metrics (C-statistics or AUC) generally ranged from moderate (≈0.70) to excellent (>0.90), with post-screening machine-learning and deep-learning approaches achieving the highest values. Calibration, when formally evaluated, was typically good for regression-based models but underreported for many complex algorithms. External validation was inconsistently applied. Available results suggest attenuation and discrimination, which is a typical manifestation of the model being overfitted in their original sample, as well as drift in calibration. The latter can also be a manifestation of overfitting, as well as differences across populations and sampling methods (e.g. trial-based data for development, and electronic health records for validation).

Screen-selection models without biomarkers were trained on routine demographic and smoking variables; most used classical Cox or logistic regression, few used non-image ML methods (e.g., gradient-boosted trees, random forests), and only one employed neural networks. In models incorporating biomarkers, inputs typically included laboratory, protein, genetic, or multi-omic markers; most used regression, some used ML, and CNNs were not used. For nodule-malignancy models, only a small subset supplement CT features with blood biomarkers, whereas the majority combine imaging with basic clinical data; approaches span regression, non-CNN ML, and CT-based CNNs.

Our meta-regression showed in pre–screening models, larger sample sizes were associated with modest but statistically significant improvements in discrimination, while the use of machine-learning algorithms, external validation, and biomarker inclusion showed favorable point estimates but did not achieve statistical significance. In post–screening models, although the baseline discrimination was substantially higher, no model feature, including sample size or advanced algorithmic approaches, emerged as a significant predictor of performance. For time-to-event data, AUCs are time-dependent and were reported inconsistently across studies, harmonization and pooling were not feasible, thus limiting our meta-regression.

These findings suggest that while sample size plays a role in model discrimination early in the screening process, other factors such as model complexity and validation strategy may have more nuanced effects. The effect size**s** in absolute terms were small (albeit this is typical for the c-statistic^93^), and the confidence bounds were wide. These findings can be used in hypothesis-generation. They may point toward patterns worth testing in future research or larger datasets, but are not confirmatory evidence of an effect. These models are developed across diverse populations: in ever-smokers and never-smokers, across Asian and Western cohorts, and in both community screenings and clinical trial settings.

Prospective cohort designs enhanced the generalizability and statistical power of several models. However, fewer than half of the models in each category underwent robust external validation, and many relied on retrospective or case–control designs that are prone to selection bias. Calibration reporting was uneven, with many machine-learning and deep-learning studies omitting calibration entirely.

With 68 new models since 2020, a substantial increase, efforts should shift from developing additional algorithms to validating those already available. Performing head-to-head comparisons of regression, machine-learning, and deep-learning approaches within the same cohorts can elucidate not only differences in accuracy, but also ease of use and acceptance by individuals, providers, and communities. In addition, future studies can assess cost-effectiveness by examining how biomarkers and imaging algorithms influence resource use, program costs, and patient outcomes. Finally, carrying out implementation studies to explore how these tools integrate into screening settings, and developing strategies for sharing individualized risk with patients. These steps will help move lung cancer risk models from development into practice.

Our review was limited by heterogeneity in model reporting: differing follow-up horizons, risk thresholds, and performance metrics complicated direct comparisons. Publication bias may favor models with higher apparent performance. Finally, rapidly evolving machine-learning methods underscore the need for continual evidence synthesis.

## Conclusions

A wide spectrum of lung cancer risk models now exists, from simple equations to sophisticated biomarker- and image-based algorithms, and with methodology varying from conventional risk equations to sophisticated machine learning algorithms. While many tools seem to demonstrate strong discrimination, the relative paucity of rigorous external validation and formal calibration reporting hampers clinical adoption. Future efforts should emphasize transparent, standardized evaluation across populations, economic evaluations, and implementation studies to realize the potential of personalized risk prediction in lung cancer screening and nodule management.

## Data Availability

All data produced in the present work are contained in the manuscript

## Supplementary Material

**Figure S1.**
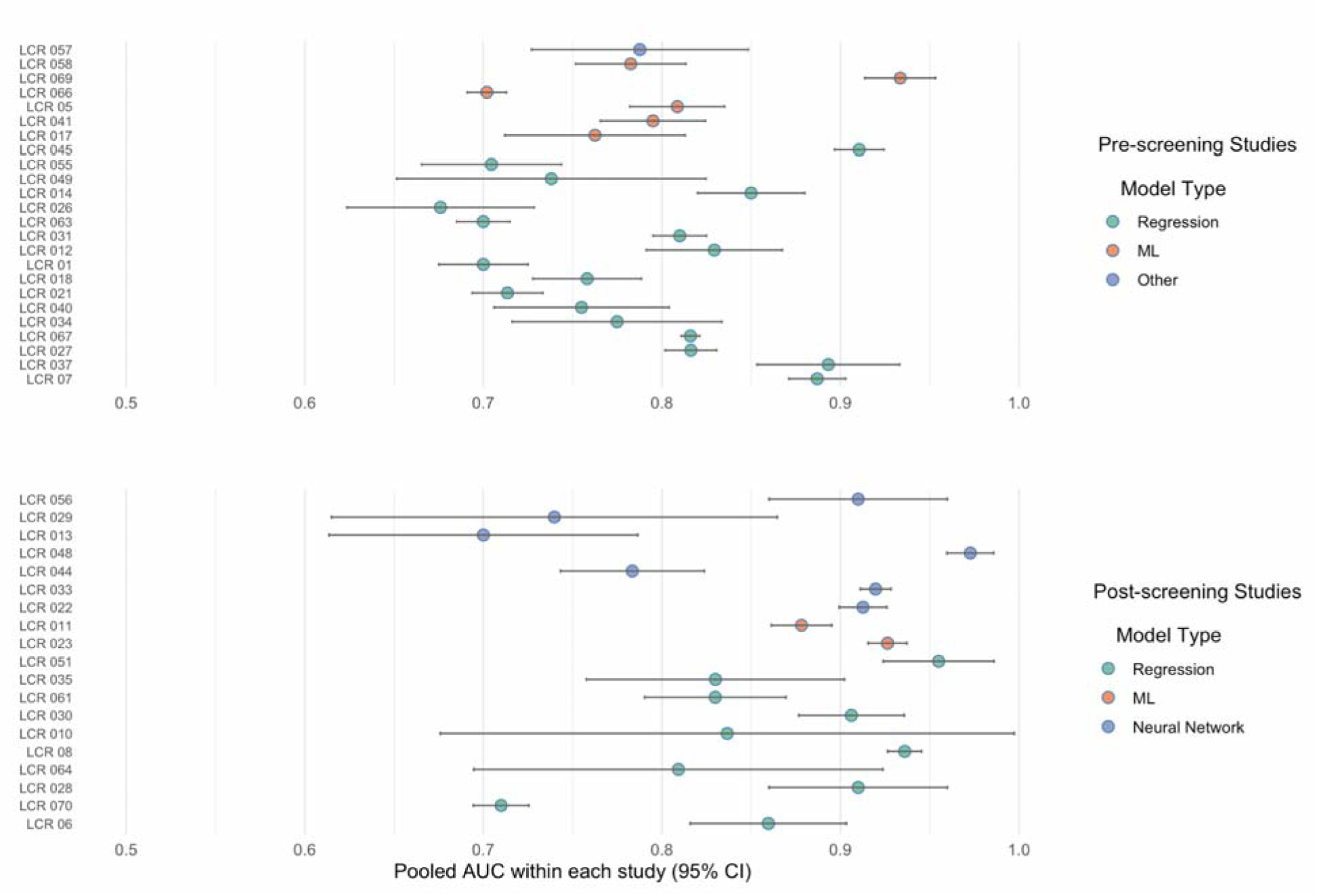
Forest plot of study-level pooled AUCs (95 % CI) by model type and screening stage

---

**Table S1:**
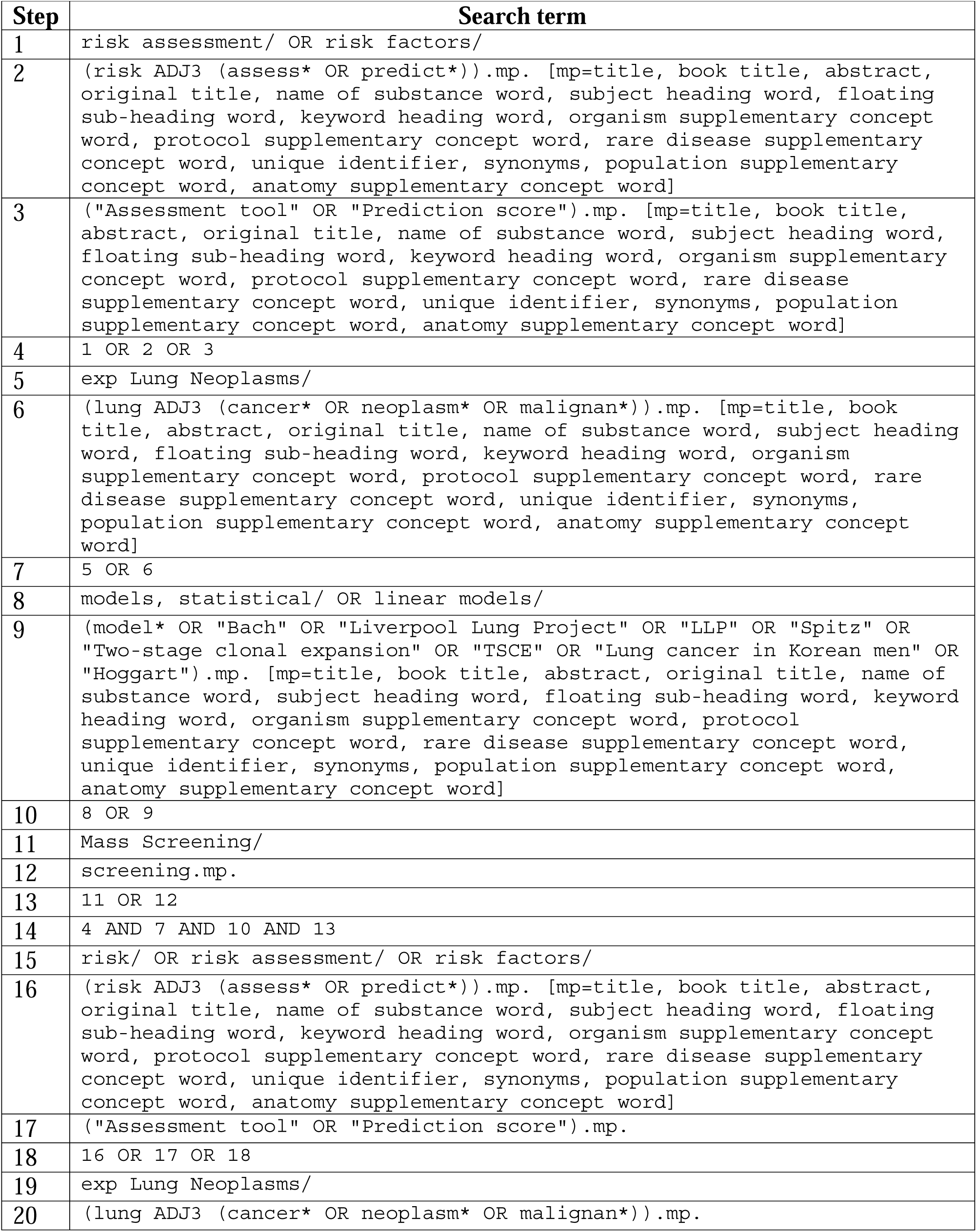
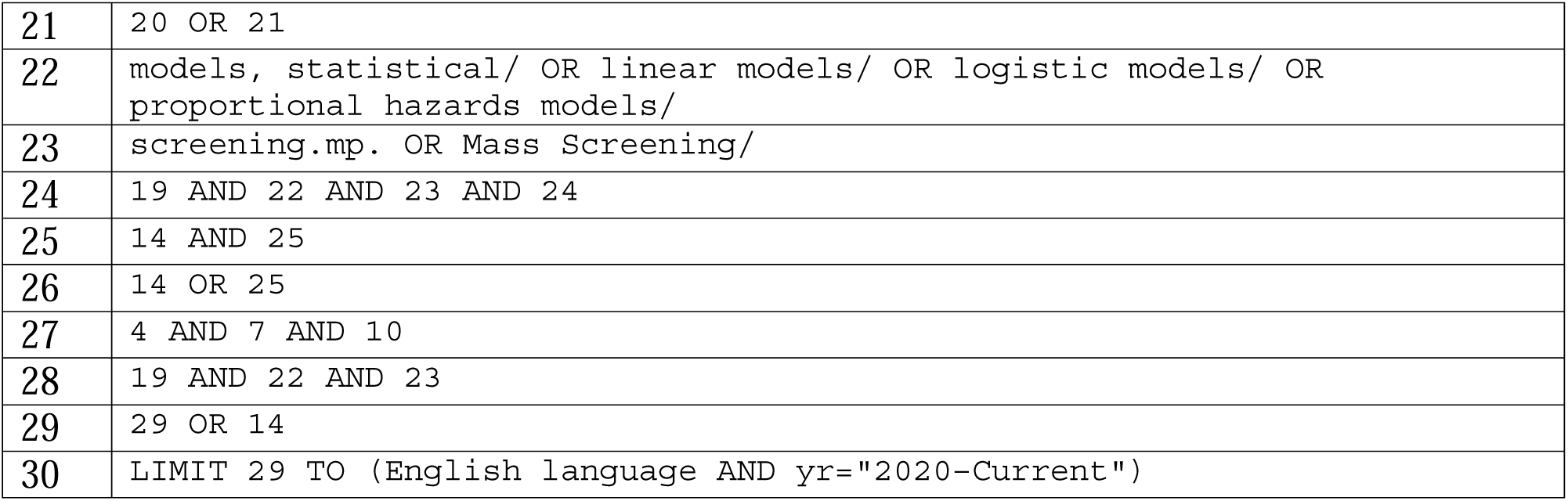
Search strategy.

| Table S1 : Search strategy |  |
| --- | --- |
| Step | Search term |
| 1 | risk assessment/ OR risk factors/ |
| 2 | (risk ADJ3 (assess* OR predict*)).mp. [mp=title, book title, abstract, original title, name of substance word, subject heading word, floating sub-heading word, keyword heading word, organism supplementary concept word, protocol supplementary concept word, rare disease supplementary concept word, unique identifier, synonyms, population supplementary concept word, anatomy supplementary concept word] |
| 3 | ("Assessment tool" OR "Prediction score").mp. [mp=title, book title, abstract, original title, name of substance word, subject heading word, floating sub-heading word, keyword heading word, organism supplementary concept word, protocol supplementary concept word, rare disease supplementary concept word, unique identifier, synonyms, population supplementary concept word, anatomy supplementary concept word] |
| 4 | 1 OR 2 OR 3 |
| 5 | exp Lung Neoplasms/ |
| 6 | (lung ADJ3 (cancer* OR neoplasm* OR malignan*)).mp. [mp=title, book title, abstract, original title, name of substance word, subject heading word, floating sub-heading word, keyword heading word, organism supplementary concept word, protocol supplementary concept word, rare disease supplementary concept word, unique identifier, synonyms, population supplementary concept word, anatomy supplementary concept word] |
| 7 | 5 OR 6 |
| 8 | models, statistical/ OR linear models/ |
| 9 | (model* OR "Bach" OR "Liverpool Lung Project" OR "LLP" OR "Spitz" OR "Two-stage clonal expansion" OR "TSCE" OR "Lung cancer in Korean men" OR "Hoggart").mp. [mp=title, book title, abstract, original title, name of substance word, subject heading word, floating sub-heading word, keyword heading word, organism supplementary concept word, protocol supplementary concept word, rare disease supplementary concept word, unique identifier, synonyms, population supplementary concept word, anatomy supplementary concept word] |
| 10 | 8 OR 9 |
| 11 | Mass Screening/ |
| 12 | screening.mp. |
| 13 | 11 OR 12 |
| 14 | 4 AND 7 AND 10 AND 13 |
| 15 | risk/ OR risk assessment/ OR risk factors/ |
| 16 | (risk ADJ3 (assess* OR predict*)).mp. [mp=title, book title, abstract, original title, name of substance word, subject heading word, floating sub-heading word, keyword heading word, organism supplementary concept word, protocol supplementary concept word, rare disease supplementary concept word, unique identifier, synonyms, population supplementary concept word, anatomy supplementary concept word] |
| 17 | ("Assessment tool" OR "Prediction score").mp. |
| 18 | 16 OR 17 OR 18 |
| 19 | exp Lung Neoplasms/ |
| 20 | (lung ADJ3 (cancer* OR neoplasm* OR malignan*)).mp. |
| 21 | 20 OR 21 |
| 22 | models, statistical/ OR linear models/ OR logistic models/ OR proportional hazards models/ |
| 23 | screening.mp. OR Mass Screening/ |
| 24 | 19 AND 22 AND 23 AND 24 |
| 25 | 14 AND 25 |
| 26 | 14 OR 25 |
| 27 | 4 AND 7 AND 10 |
| 28 | 19 AND 22 AND 23 |
| 29 | 29 OR 14 |
| 30 | LIMIT 29 TO (English language AND yr="2020-Current") |

**Table S2.** RVE Hierarchical Effects Model (Pre-screening)

| Table S2. RVE Hierarchical Effects Model (Pre-screening) |  |  |  |  |  |  |  |
| --- | --- | --- | --- | --- | --- | --- | --- |
| Variable | Estimate | Std. Error | df | P-value | 95% CI Lower | 95% CI Upper | Sig. |
| (Intercept) | 0.74214 | 0.02582 | 7.81 | 0.0000000033 | 0.68234 | 0.80194 | *** |
| √Sample Size | 0.0000756 | 0.0000226 | 3.91 | 0.02973 | 0.0000123 | 0.000139 | ** |
| Model Category: ML | 0.03711 | 0.03414 | 8.44 | 0.30705 | −0.04091 | 0.11513 |  |
| Model Category: Other | −0.00552 | 0.03330 | 11.71 | 0.87106 | −0.07827 | 0.06722 |  |
| Validation Type: external | −0.01158 | 0.01403 | 6.39 | 0.43887 | −0.04541 | 0.02225 |  |
| Screening Stage: prescreen (BM) | 0.03322 | 0.03316 | 11.60 | 0.33693 | −0.03931 | 0.10575 |  |

**Table S3.** RVE Hierarchical Effects Model (Post-screening)

| Table S3. RVE Hierarchical Effects Model (Post-screening) |  |  |  |  |  |  |  |
| --- | --- | --- | --- | --- | --- | --- | --- |
| Variable | Estimate | Std. Error | df | P-value | 95% CI Lower | 95% CI Upper | Sig. |
| (Intercept) | 0.87993 | 0.02398 | 5.33 | 0.00000013 | 0.81942 | 0.94045 | *** |
| √Sample Size | −0.000108 | 0.000335 | 5.21 | 0.75863 | −0.000959 | 0.000742 |  |
| Model Category: ML | 0.03492 | 0.03982 | 1.94 | 0.47557 | −0.14190 | 0.21174 |  |
| Model Category: Neural Network | 0.04712 | 0.05414 | 6.52 | 0.41497 | −0.08282 | 0.17707 |  |
| Validation Type: external | −0.00133 | 0.02397 | 7.06 | 0.95724 | −0.05792 | 0.05525 |  |

## References

1. Bray F, Ferlay J, Soerjomataram I, Siegel RL, Torre LA, Jemal A. Global cancer statistics 2018: GLOBOCAN estimates of incidence and mortality worldwide for 36 cancers in 185 countries. CA: A Cancer Journal for Clinicians. 2018;68(6):394–424. doi:10.3322/caac.21492

2. cancer CCS/ S canadienne du. Survival statistics for non–small cell lung cancer. Canadian Cancer Society. August 2023. Accessed April 8, 2025. https://cancer.ca/en/cancer-information/cancer-types/lung/prognosis-and-survival/non-small-cell-lung-cancer-survival-statistics

3. CDC. U.S. Cancer Statistics Lung Cancer Stat Bite. United States Cancer Statistics. June 5, 2024. Accessed May 22, 2025. https://www.cdc.gov/united-states-cancer-statistics/publications/lung-cancer-stat-bite.html

4. Nations JA, Brown DW, Shao S, Shriver CD, Zhu K. Comparative Trends in the Distribution of Lung Cancer Stage at Diagnosis in the Department of Defense Cancer Registry and the Surveillance, Epidemiology, and End Results data, 1989-2012. Mil Med. 2020;185(11-12):e2044-e2048. doi:10.1093/milmed/usaa218

5. National Lung Screening Trial Research Team, Aberle DR, Adams AM, et al. Reduced lung-cancer mortality with low-dose computed tomographic screening. N Engl J Med. 2011;365(5):395–409. doi:10.1056/NEJMoa1102873

6. The National Lung Screening Trial: Overview and Study Design1. Radiology. 2011;258(1):243–253. doi:10.1148/radiol.10091808

7. Wille MMW, Dirksen A, Ashraf H, et al. Results of the Randomized Danish Lung Cancer Screening Trial with Focus on High-Risk Profiling. Am J Respir Crit Care Med. 2016;193(5):542–551. doi:10.1164/rccm.201505-1040OC

8. Pastorino U, Rossi M, Rosato V, et al. Annual or biennial CT screening versus observation in heavy smokers: 5-year results of the MILD trial. Eur J Cancer Prev. 2012;21(3):308–315. doi:10.1097/CEJ.0b013e328351e1b6

9. Paci E, Puliti D, Lopes Pegna A, et al. Mortality, survival and incidence rates in the ITALUNG randomised lung cancer screening trial. Thorax. 2017;72(9):825–831. doi:10.1136/thoraxjnl-2016-209825

10. Infante M, Cavuto S, Lutman FR, et al. Long-Term Follow-up Results of the DANTE Trial, a Randomized Study of Lung Cancer Screening with Spiral Computed Tomography. Am J Respir Crit Care Med. 2015;191(10):1166–1175. doi:10.1164/rccm.201408-1475OC

11. Horeweg N, van der Aalst CM, Vliegenthart R, et al. Volumetric computed tomography screening for lung cancer: three rounds of the NELSON trial. Eur Respir J. 2013;42(6):1659–1667. doi:10.1183/09031936.00197712

12. Gohagan JK, Marcus PM, Fagerstrom RM, et al. Final results of the Lung Screening Study, a randomized feasibility study of spiral CT versus chest X-ray screening for lung cancer. Lung Cancer. 2005;47(1):9–15. doi:10.1016/j.lungcan.2004.06.007

13. Blanchon T, Bréchot JM, Grenier PA, et al. Baseline results of the Depiscan study: a French randomized pilot trial of lung cancer screening comparing low dose CT scan (LDCT) and chest X-ray (CXR). Lung Cancer. 2007;58(1):50–58. doi:10.1016/j.lungcan.2007.05.009

14. null null. Reduced Lung-Cancer Mortality with Low-Dose Computed Tomographic Screening. New England Journal of Medicine. 2011;365(5):395–409. doi:10.1056/NEJMoa1102873

15. Darden ME, Hoagland A. Lung Cancer Screening and USPSTF Recommendations. JAMA Network Open. 2025;8(2):e2458916. doi:10.1001/jamanetworkopen.2024.58916

16. Arnold H. Paving the way for lung cancer screening: learning from Central and Eastern Europe. The Lung Cancer Policy Network. October 2, 2024. Accessed April 21, 2025. https://www.lungcancerpolicynetwork.com/paving-the-way-for-lung-cancer-screening-learning-from-central-and-eastern-europe/

17. Borilova S, Dusek L, Jakubikova L, et al. Lung Cancer in the Czech Republic. Journal of Thoracic Oncology. 2023;18(3):271–277. doi:10.1016/j.jtho.2022.11.023

18. Wait S, Alvarez-Rosete A, Osama T, et al. Implementing Lung Cancer Screening in Europe: Taking a Systems Approach. JTO Clin Res Rep. 2022;3(5):100329. doi:10.1016/j.jtocrr.2022.100329

19. Ward B, Vašáková MK, Cordeiro CR, et al. Important steps towards a big change for lung health: a joint approach by the European Respiratory Society, the European Society of Radiology and their partners to facilitate implementation of the European Union’s new recommendations on lung cancer screening. ERJ Open Research. 2023;9(3). doi:10.1183/23120541.00026-2023

20. Care CTF on PH. Recommendations on screening for lung cancer. CMAJ. 2016;188(6):425–432. doi:10.1503/cmaj.151421

21. Pinsky PF, Berg CD. Applying the National Lung Screening Trial eligibility criteria to the US population: what percent of the population and of incident lung cancers would be covered? J Med Screen. 2012;19(3):154–156. doi:10.1258/jms.2012.012010

22. Gracie K, Kennedy MPT, Esterbrook G, et al. The proportion of lung cancer patients attending UK lung cancer clinics who would have been eligible for low-dose CT screening. Eur Respir J. 2019;54(2):1802221. doi:10.1183/13993003.02221-2018

23. Toumazis I, Bastani M, Han SS, Plevritis SK. Risk-Based lung cancer screening: A systematic review. Lung Cancer. 2020;147:154–186. doi:10.1016/j.lungcan.2020.07.007

24. Fernandez-Felix BM, López-Alcalde J, Roqué M, Muriel A, Zamora J. CHARMS and PROBAST at your fingertips: a template for data extraction and risk of bias assessment in systematic reviews of predictive models. BMC Medical Research Methodology. 2023;23(1):44. doi:10.1186/s12874-023-01849-0

25. Yang JJ, Wen W, Zahed H, et al. Lung Cancer Risk Prediction Models for Asian Ever-Smokers. J Thorac Oncol. 2024;19(3):451–464. doi:10.1016/j.jtho.2023.11.002

26. Levi M, Lazebnik T, Kushnir S, Yosef N, Shlomi D. Machine learning computational model to predict lung cancer using electronic medical records. Cancer Epidemiol. 2024;92:102631. doi:10.1016/j.canep.2024.102631

27. Guo LW, Lyu ZY, Meng QC, et al. A risk prediction model for selecting high-risk population for computed tomography lung cancer screening in China. Lung Cancer. 2022;163:27–34. doi:10.1016/j.lungcan.2021.11.015

28. Pan Z, Zhang R, Shen S, et al. OWL: an optimized and independently validated machine learning prediction model for lung cancer screening based on the UK Biobank, PLCO, and NLST populations. EBioMedicine. 2023;88:104443. doi:10.1016/j.ebiom.2023.104443

29. Liao W, Coupland CAC, Burchardt J, et al. Predicting the future risk of lung cancer: development, and internal and external validation of the CanPredict (lung) model in 19·67 million people and evaluation of model performance against seven other risk prediction models. The Lancet Respiratory Medicine. 2023;11(8):685–697. doi:10.1016/S2213-2600(23)00050-4

30. Guo L, Meng Q, Zheng L, et al. Lung Cancer Risk Prediction Nomogram in Nonsmoking Chinese Women: Retrospective Cross-sectional Cohort Study. JMIR Public Health Surveill. 2023;9:e41640. doi:10.2196/41640

31. Howell D, Buttery R, Badrinath P, et al. Developing a risk prediction tool for lung cancer in Kent and Medway, England: cohort study using linked data. BJC Rep. 2023;1(1):16. doi:10.1038/s44276-023-00019-5

32. Chandran U, Reps J, Yang R, Vachani A, Maldonado F, Kalsekar I. Machine Learning and Real-World Data to Predict Lung Cancer Risk in Routine Care. Cancer Epidemiol Biomarkers Prev. 2023;32(3):337–343. doi:10.1158/1055-9965.EPI-22-0873

33. Ma Z, Lv J, Zhu M, et al. Lung cancer risk score for ever and never smokers in China. Cancer Commun (Lond). 2023;43(8):877–895. doi:10.1002/cac2.12463

34. Wang F, Tan F, Shen S, et al. Risk-stratified Approach for Never- and Ever-Smokers in Lung Cancer Screening: A Prospective Cohort Study in China. Am J Respir Crit Care Med. 2023;207(1):77–88. doi:10.1164/rccm.202204-0727OC

35. Yeo Y, Shin DW, Han K, et al. Individual 5-Year Lung Cancer Risk Prediction Model in Korea Using a Nationwide Representative Database. Cancers (Basel). 2021;13(14):3496. doi:10.3390/cancers13143496

36. Field JK, Vulkan D, Davies MPA, Duffy SW, Gabe R. Liverpool Lung Project lung cancer risk stratification model: calibration and prospective validation. Thorax. 2021;76(2):161–168. doi:10.1136/thoraxjnl-2020-215158

37. Jani BD, Sullivan MK, Hanlon P, et al. Personalised lung cancer risk stratification and lung cancer screening: do general practice electronic medical records have a role? Br J Cancer. 2023;129(12):1968–1977. doi:10.1038/s41416-023-02467-9

38. Chien LH, Chen TY, Chen CH, et al. Recalibrating Risk Prediction Models by Synthesizing Data Sources: Adapting the Lung Cancer PLCO Model for Taiwan. Cancer Epidemiol Biomarkers Prev. 2022;31(12):2208–2218. doi:10.1158/1055-9965.EPI-22-0281

39. Jung AW, Holm PC, Gaurav K, et al. Multi-cancer risk stratification based on national health data: a retrospective modelling and validation study. The Lancet Digital Health. 2024;6(6):e396–e406. doi:10.1016/S2589-7500(24)00062-1

40. Callender T, Imrie F, Cebere B, et al. Assessing eligibility for lung cancer screening using parsimonious ensemble machine learning models: A development and validation study. PLOS Medicine. 2023;20(10):e1004287. doi:10.1371/journal.pmed.1004287

41. Yeh MCH, Wang YH, Yang HC, Bai KJ, Wang HH, Li YCJ. Artificial Intelligence-Based Prediction of Lung Cancer Risk Using Nonimaging Electronic Medical Records: Deep Learning Approach. J Med Internet Res. 2021;23(8):e26256. doi:10.2196/26256

42. Guo LW, Lyu ZY, Meng QC, et al. Construction and Validation of a Lung Cancer Risk Prediction Model for Non-Smokers in China. Front Oncol. 2022;11:766939. doi:10.3389/fonc.2021.766939

43. Alonso E, Calle X, Gurrutxaga I, Beristain A. Survival Stacking Ensemble Model for Lung Cancer Risk Prediction. Stud Health Technol Inform. 2024;321:155–159. doi:10.3233/SHTI241083

44. Park B, Kim Y, Lee J, Lee N, Jang SH. Risk-based prediction model for selecting eligible population for lung cancer screening among ever smokers in Korea. Transl Lung Cancer Res. 2021;10(12):4390–4402. doi:10.21037/tlcr-21-566

45. Gould MK, Huang BZ, Tammemagi MC, Kinar Y, Shiff R. Machine Learning for Early Lung Cancer Identification Using Routine Clinical and Laboratory Data. Am J Respir Crit Care Med. 2021;204(4):445–453. doi:10.1164/rccm.202007-2791OC

46. Fahrmann JF, Marsh T, Irajizad E, et al. Blood-Based Biomarker Panel for Personalized Lung Cancer Risk Assessment. J Clin Oncol. 2022;40(8):876–883. doi:10.1200/JCO.21.01460

47. Zhang S, Yang L, Xu W, et al. Predicting the risk of lung cancer using machine learning: A large study based on UK Biobank. Medicine (Baltimore). 2024;103(16):e37879. doi:10.1097/MD.0000000000037879

48. Ma Z, Zhu Z, Pang G, et al. Development and validation of a lung cancer polygenic risk score incorporating susceptibility variants for risk factors. Int J Cancer. 2025;156(5):953–963. doi:10.1002/ijc.35210

49. Cortés-Ibáñez FO, Johnson T, Mascalchi M, Katzke V, Delorme S, Kaaks R. Serum-based biomarkers associated with lung cancer risk and cause-specific mortality in the German randomized Lung Cancer Screening Intervention (LUSI) trial. Transl Lung Cancer Res. 2023;12(12):2460–2475. doi:10.21037/tlcr-23-548

50. Horsfall LJ, Clarke CS, Nazareth I, Ambler G. The value of blood-based measures of liver function and urate in lung cancer risk prediction: A cohort study and health economic analysis. Cancer Epidemiol. 2023;84:102354. doi:10.1016/j.canep.2023.102354

51. Nguyen OTD, Fotopoulos I, Nøst TH, et al. The HUNT lung-SNP model: genetic variants plus clinical variables improve lung cancer risk assessment over clinical models. J Cancer Res Clin Oncol. 2024;150(8):389. doi:10.1007/s00432-024-05909-w

52. Chen A, Wu E, Huang R, et al. Development of Lung Cancer Risk Prediction Machine Learning Models for Equitable Learning Health System: Retrospective Study. JMIR AI. 2024;3:e56590. doi:10.2196/56590

53. Wang Z, Xie K, Zhu G, et al. Early detection and stratification of lung cancer aided by a cost-effective assay targeting circulating tumor DNA (ctDNA) methylation. Respir Res. 2023;24(1):163. doi:10.1186/s12931-023-02449-8

54. Yu H, Raut JR, Schöttker B, Holleczek B, Zhang Y, Brenner H. Individual and joint contributions of genetic and methylation risk scores for enhancing lung cancer risk stratification: data from a population-based cohort in Germany. Clinical Epigenetics. 2020;12(1):89. doi:10.1186/s13148-020-00872-y

55. Tang Y, You D, Yi H, Yang S, Zhao Y. IPRS: Leveraging Gene-Environment Interaction to Reconstruct Polygenic Risk Score. Front Genet. 2022;13:801397. doi:10.3389/fgene.2022.801397

56. Chien LH, Chen CH, Chen TY, et al. Predicting Lung Cancer Occurrence in Never-Smoking Females in Asia: TNSF-SQ, a Prediction Model. Cancer Epidemiol Biomarkers Prev. 2020;29(2):452–459. doi:10.1158/1055-9965.EPI-19-1221

57. Hung RJ, Warkentin MT, Brhane Y, et al. Assessing Lung Cancer Absolute Risk Trajectory Based on a Polygenic Risk Model. Cancer Res. 2021;81(6):1607–1615. doi:10.1158/0008-5472.CAN-20-1237

58. Bhardwaj M, Schöttker B, Holleczek B, Benner A, Schrotz-King P, Brenner H. Potential of Inflammatory Protein Signatures for Enhanced Selection of People for Lung Cancer Screening. Cancers (Basel). 2022;14(9):2146. doi:10.3390/cancers14092146

59. Zhang S, Yang F, Wang L, Si S, Zhang J, Xue F. Personalized prediction for multiple chronic diseases by developing the multi-task Cox learning model. PLoS Comput Biol. 2023;19(9):e1011396. doi:10.1371/journal.pcbi.1011396

60. Jia G, Lu Y, Wen W, et al. Evaluating the Utility of Polygenic Risk Scores in Identifying High-Risk Individuals for Eight Common Cancers. JNCI Cancer Spectr. 2020;4(3):pkaa021. doi:10.1093/jncics/pkaa021

61. Jacobsen KK, Kobylecki CJ, Skov-Jeppesen SM, Bojesen SE. Development and validation of a simple general population lung cancer risk model including AHRR-methylation. Lung Cancer. 2023;181:107229. doi:10.1016/j.lungcan.2023.107229

62. Lian J, Vardhanabhuti V. Metabolic biomarkers using nuclear magnetic resonance metabolomics assay for the prediction of aging-related disease risk and mortality: a prospective, longitudinal, observational, cohort study based on the UK Biobank. Geroscience. 2024;46(2):1515–1526. doi:10.1007/s11357-023-00918-y

63. Wei X, Sun D, Gao J, et al. Development and evaluation of a polygenic risk score for lung cancer in never-smoking women: A large-scale prospective Chinese cohort study. Int J Cancer. 2024;154(5):807–815. doi:10.1002/ijc.34765

64. Yao Y, Wang X, Guan J, et al. Metabolomic differentiation of benign vs malignant pulmonary nodules with high specificity via high-resolution mass spectrometry analysis of patient sera. Nat Commun. 2023;14(1):2339. doi:10.1038/s41467-023-37875-1

65. Wang N, Yao C, Luo C, et al. Integrated plasma and exosome long noncoding RNA profiling is promising for diagnosing non-small cell lung cancer. Clinical Chemistry and Laboratory Medicine (CCLM). 2023;61(12):2216–2228. doi:10.1515/cclm-2023-0291

66. Schreuder A, Jacobs C, Lessmann N, et al. Combining pulmonary and cardiac computed tomography biomarkers for disease-specific risk modelling in lung cancer screening. Eur Respir J. 2021;58(3):2003386. doi:10.1183/13993003.03386-2020

67. Chetan MR, Dowson N, Price NW, Ather S, Nicolson A, Gleeson FV. Developing an understanding of artificial intelligence lung nodule risk prediction using insights from the Brock model. Eur Radiol. 2022;32(8):5330–5338. doi:10.1007/s00330-022-08635-4

68. Yuan H, Gao Z, He X, et al. Application of logistic regression and convolutional neural network in prediction and diagnosis of high-risk populations of lung cancer. Eur J Cancer Prev. 2022;31(2):145–151. doi:10.1097/CEJ.0000000000000684

69. Wang Y, Zhou C, Ying L, et al. Leveraging Serial Low-Dose CT Scans in Radiomics-based Reinforcement Learning to Improve Early Diagnosis of Lung Cancer at Baseline Screening. Radiol Cardiothorac Imaging. 2024;6(3):e230196. doi:10.1148/ryct.230196

70. Lu MT, Raghu VK, Mayrhofer T, Aerts HJWL, Hoffmann U. Deep Learning Using Chest Radiographs to Identify High-Risk Smokers for Lung Cancer Screening Computed Tomography: Development and Validation of a Prediction Model. Ann Intern Med. 2020;173(9):704–713. doi:10.7326/M20-1868

71. Choi K, Park JS, Kwon YS, et al. Development of lung cancer risk prediction models based on F-18 FDG PET images. Ann Nucl Med. 2023;37(10):572–582. doi:10.1007/s12149-023-01858-5

72. Chang HT, Wang PH, Chen WF, Lin CJ. Risk Assessment of Early Lung Cancer with LDCT and Health Examinations. Int J Environ Res Public Health. 2022;19(8):4633. doi:10.3390/ijerph19084633

73. Venkadesh KV, Setio AAA, Schreuder A, et al. Deep Learning for Malignancy Risk Estimation of Pulmonary Nodules Detected at Low-Dose Screening CT. Radiology. 2021;300(2):438–447. doi:10.1148/radiol.2021204433

74. Warkentin MT, Al-Sawaihey H, Lam S, et al. Radiomics analysis to predict pulmonary nodule malignancy using machine learning approaches. Thorax. 2024;79(4):307–315. doi:10.1136/thorax-2023-220226

75. Yao B, Huang X, Wu F, et al. A Novel Model Using Serum Thymidine Kinase 1 and Low-dose Computed Tomography Parameters to Predict Three-year Lung Cancer Risk in People with Pulmonary Nodules: A Retrospective Study. J Cancer. 2024;15(3):737–746. doi:10.7150/jca.90428

76. Meng Q, Li B, Gao P, et al. Development and Validation of a Risk Stratification Model of Pulmonary Ground-Glass Nodules Based on Complementary Lung-RADS 1.1 and Deep Learning Scores. Front Public Health. 2022;10:891306. doi:10.3389/fpubh.2022.891306

77. Liu D, Sun X, Liu A, et al. Predictive value of a novel Asian lung cancer screening nomogram based on artificial intelligence and epidemiological characteristics. Thorac Cancer. 2021;12(23):3130–3140. doi:10.1111/1759-7714.14140

78. Massion PP, Antic S, Ather S, et al. Assessing the Accuracy of a Deep Learning Method to Risk Stratify Indeterminate Pulmonary Nodules. Am J Respir Crit Care Med. 2020;202(2):241–249. doi:10.1164/rccm.201903-0505OC

79. Lu H, Kim J, Qi J, et al. Multi-Window CT Based Radiological Traits for Improving Early Detection in Lung Cancer Screening. Cancer Manag Res. 2020;12:12225–12238. doi:10.2147/CMAR.S246609

80. Zhou C, Chan HP, Chughtai A, Hadjiiski LM, Kazerooni EA, Wei J. Pathologic categorization of lung nodules: Radiomic descriptors of CT attenuation distribution patterns of solid and subsolid nodules in low-dose CT. Eur J Radiol. 2020;129:109106. doi:10.1016/j.ejrad.2020.109106

81. Rundo L, Ledda RE, di Noia C, et al. A Low-Dose CT-Based Radiomic Model to Improve Characterization and Screening Recall Intervals of Indeterminate Prevalent Pulmonary Nodules. Diagnostics (Basel). 2021;11(9):1610. doi:10.3390/diagnostics11091610

82. Mikhael PG, Wohlwend J, Yala A, et al. Sybil: A Validated Deep Learning Model to Predict Future Lung Cancer Risk From a Single Low-Dose Chest Computed Tomography. JCO. 2023;41(12):2191–2200. doi:10.1200/JCO.22.01345

83. Venkadesh KV, Aleef TA, Scholten ET, et al. Prior CT Improves Deep Learning for Malignancy Risk Estimation of Screening-detected Pulmonary Nodules. Radiology. 2023;308(2):e223308. doi:10.1148/radiol.223308

84. Liu J, Xu H, Qing H, et al. Comparison of Radiomic Models Based on Low-Dose and Standard-Dose CT for Prediction of Adenocarcinomas and Benign Lesions in Solid Pulmonary Nodules. Front Oncol. 2021;10:634298. doi:10.3389/fonc.2020.634298

85. Ebrahimpour L, Després P, Manem VSK. Differential Radiomics-Based Signature Predicts Lung Cancer Risk Accounting for Imaging Parameters in NLST Cohort. Cancer Med. 2024;13(20):e70359. doi:10.1002/cam4.70359

86. Gao R, Tang Y, Khan MS, et al. Cancer Risk Estimation Combining Lung Screening CT with Clinical Data Elements. Radiol Artif Intell. 2021;3(6):e210032. doi:10.1148/ryai.2021210032

87. Xiao H, Shi Z, Zou Y, et al. One-off low-dose CT screening of positive nodules in lung cancer: A prospective community-based cohort study. Lung Cancer. 2023;177:1–10. doi:10.1016/j.lungcan.2023.01.005

88. Wu Z, Tan F, Xie Y, et al. A strategy to reduce the false-positive rate after low-dose computed tomography in lung cancer screening: A multicenter prospective cohort study. Cancer Med. 2023;12(13):14781–14793. doi:10.1002/cam4.6106

89. Xu K, Khan MS, Li TZ, et al. AI Body Composition in Lung Cancer Screening: Added Value Beyond Lung Cancer Detection. Radiology. 2023;308(1):e222937. doi:10.1148/radiol.222937

90. Trajanovski S, Mavroeidis D, Swisher CL, et al. Towards radiologist-level cancer risk assessment in CT lung screening using deep learning. Comput Med Imaging Graph. 2021;90:101883. doi:10.1016/j.compmedimag.2021.101883

91. Paul R, Schabath MB, Gillies R, Hall LO, Goldgof DB. Hybrid models for lung nodule malignancy prediction utilizing convolutional neural network ensembles and clinical data. J Med Imaging (Bellingham). 2020;7(2):024502. doi:10.1117/1.JMI.7.2.024502

92. Robbins HA, Cheung LC, Chaturvedi AK, Baldwin DR, Berg CD, Katki HA. Management of Lung Cancer Screening Results Based on Individual Prediction of Current and Future Lung Cancer Risks. Journal of Thoracic Oncology. 2022;17(2):252–263. doi:10.1016/j.jtho.2021.10.001

93. Baker SG, Schuit E, Steyerberg EW, et al. How to interpret a small increase in AUC with an additional risk prediction marker: Decision analysis comes through. Stat Med. 2014;33(22):3946–3959. doi:10.1002/sim.6195

